# TMS-induced plasticity improving cognitive control in OCD I: Clinical and neuroimaging outcomes from a randomised trial of rTMS for OCD

**DOI:** 10.1101/2023.11.04.23298100

**Authors:** Sophie M.D.D. Fitzsimmons, Tjardo Postma, A. Dilene van Campen, Chris Vriend, Neeltje M. Batelaan, Patricia van Oppen, Adriaan W. Hoogendoorn, Ysbrand D. van der Werf, Odile A. van den Heuvel

**Affiliations:** Amsterdam UMC, Vrije Universiteit Amsterdam, Department of Psychiatry, de Boelelaan 1117, Amsterdam, The Netherlands; Amsterdam UMC, Vrije Universiteit Amsterdam, Department of Anatomy & Neurosciences, de Boelelaan 1117, Amsterdam, The Netherlands; Amsterdam Neuroscience, Compulsivity, Impulsivity & Attention program, Amsterdam, The Netherlands; Amsterdam Public Health, Amsterdam, The Netherlands; GGZ inGeest, Amsterdam, The Netherlands

**Keywords:** functional MRI, rTMS, OCD, cognitive control, task-based fMRI, exposure and response prevention psychotherapy

## Abstract

**Background:** Repetitive transcranial magnetic stimulation (rTMS) is an emerging treatment option for obsessive-compulsive disorder (OCD). The neurobiological mechanisms of rTMS in OCD have, however, been incompletely characterized. We compared clinical outcomes and changes in task-based brain activation following three different rTMS stimulation protocols, all combined with exposure and response prevention (ERP).

**Methods:** In this three-arm proof-of-concept randomized controlled clinical trial, 61 treatment-refractory adult OCD patients received 16 sessions of rTMS immediately prior to ERP over 8 weeks, with task-based functional MRI (tb-fMRI) scans and clinical assessments pre- and post-treatment. Patients received either: high frequency (HF) rTMS to the left dorsolateral prefrontal cortex (DLPFC) (n=19 (6M/13F)); HF rTMS to the left pre-supplementary motor area (preSMA) (n=23 (10M/13F)); or control rTMS to the vertex (n=19 (6M/13F)). Changes in tb-fMRI activation pre-post treatment were compared using both a Bayesian region-of-interest and a general linear model whole-brain approach.

**Results:** Mean OCD symptom severity decreased significantly in all treatment groups (delta=- 10.836, p<0.001, 95% CI [-12.504, -9.168]), with no differences between groups. Response rate in the entire sample was 57.4%. Groups receiving DLPFC or preSMA rTMS showed, respectively, a decrease in planning and error processing task-related activation after treatment that was associated with symptom improvement, while individuals in the vertex rTMS group with greater symptom improvement showed an increase in inhibition-related activation.

**Conclusions:** PreSMA and DLPFC rTMS combined with ERP led to significant symptom improvement related to activation decreases in targeted task networks, although we observed no differences in symptom reduction between groups.

This trial was registered at clinicaltrials.gov (NCT03667807)

## 1. Introduction

Current first-line treatments for obsessive-compulsive disorder (OCD) include cognitive-behavioral or exposure and response prevention (ERP) psychotherapy; and pharmacotherapy, usually with serotonin reuptake inhibitors (SRIs). However, these treatments are only fully effective in ∼50% of patients(1). Deep brain stimulation is an option for highly treatment-resistant patients, but due to the potential risks and side effects of this invasive procedure(2), a need exists for alternative non-invasive neuromodulation options for OCD patients who do not respond to first-line treatments.

OCD symptoms are linked to functional and structural alterations in fronto-striatal, fronto-limbic and fronto-parietal brain circuits(3). The dorsolateral prefrontal cortex (DLPFC) and pre-supplementary motor area (preSMA) are key regions of cortico-striato-thalamo-cortical (CSTC) circuits that are involved in cognitive control and affected in OCD. The DLPFC plays a role in emotion regulation and planning and shows altered activation in OCD compared to healthy controls: increased activation during working memory, and decreased activation during planning and emotion regulation(4–6). The preSMA is important for response inhibition (the ability to stop an initiated action), and shows compensatory hyperactivation in OCD patients and unaffected siblings, with greater hyperactivation associated with better response inhibition task performance(6,7).

Previous studies have shown that repetitive transcranial magnetic stimulation (rTMS) relieves OCD symptoms(8). Repetitive TMS induces plastic changes in brain circuits by modulating the activity of the stimulated region and (functionally) connected brain areas(9–12). The effects of various rTMS treatments on brain activity and their relationship to treatment outcome in OCD have, however, not yet been fully described.

To address this, we carried out a randomized proof-of-concept trial in which OCD patients resistant to first-line treatments were assigned to one of three different high-frequency rTMS conditions: left DLPFC; left preSMA; or low-intensity rTMS to the vertex as a control condition. All patients received ERP directly following each rTMS session, and underwent pre-post treatment task-based functional MRI (tb-fMRI) scans.

The aim of the present study was to characterize and compare treatment-induced changes in brain activation caused by three different combined rTMS-ERP protocols in OCD using tb-fMRI. By modulating activation in networks related to specific cognitive functions using high-frequency rTMS, we intended to enhance neuroplasticity processes that could improve the outcome of concurrent ERP. While 10Hz DLPFC stimulation has previously been used successfully in OCD, the preSMA is usually stimulated with 1Hz rTMS, classically thought to decrease the excitability of the stimulated region. We however chose to administer 10Hz rTMS, as we aimed to increase preSMA activation to emulate the compensatory increased preSMA activation seen in unaffected siblings of OCD patients during response inhibition(7).

The primary outcome measure was change in planning- and inhibition-related brain activation following treatment. We hypothesized that DLPFC (versus vertex) stimulation would increase planning-related activation in the DLPFC and other regions important for planning; and that preSMA (versus vertex) stimulation would increase inhibition-related activation in preSMA and other regions in the response inhibition network. The secondary outcome measure was change in OCD symptom severity following treatment. We hypothesized that rTMS to the DLPFC and preSMA combined with ERP would cause a greater improvement in OCD symptoms than the vertex + ERP condition. Finally, as an exploratory outcome, we examined the relationship between change in brain activation and change in symptom severity.

## 2. Methods and materials

The following methods are abbreviated; full methodological details are reported in the Supplement.

### 2.1 Participants

This trial was registered at clinicaltrials.gov (NCT03667807), and is reported according to CONSORT guidelines(13) (see Table S1 for CONSORT checklist). Sixty-six adults with OCD were randomized. Eligibility criteria were: meeting DSM-5 criteria for OCD as determined by the Structured Clinical Interview for DSM-5(14), moderate-severe OCD symptoms (Yale-Brown Obsessive-Compulsive Scale(15) (YBOCS) score of ≥16), age 18-65 years, previous treatment with ≥8 sessions of ERP or cognitive-behavioral therapy for OCD, and ≥12 weeks of previous SRI treatment for OCD or medication naïve with a strong preference for non-pharmacological treatment. Exclusion criteria were: Tourette’s syndrome, schizophrenia, bipolar disorder, active suicidal ideation, MRI/rTMS exclusion criteria, and previous rTMS treatment. The study was approved by the ethics committee of the VU Medical Center (VUmc), Amsterdam, The Netherlands, and was conducted as a collaboration between the VUmc and GGZ inGeest, Amsterdam. All participants provided written informed consent in accordance with the declaration of Helsinki.

### 2.2 Study design

This was a three-arm parallel randomized controlled trial, conducted between May 2019 and October 2022. Participants received one of three types of rTMS (targeting either L DLPFC, L preSMA, or vertex at a low stimulation intensity as a control condition), immediately followed by ERP twice a week, for 8 weeks (20 treatment sessions in total, of which 16 were rTMS-ERP sessions) (see section 2.3 for details of rTMS and ERP treatments). OCD symptom severity was assessed using the YBOCS before treatment (T0), after rTMS-ERP session 8 (T1), after rTMS-ERP session 16 (T2), and 12 weeks following the end of treatment (T3). Participants also underwent multimodal MRI scans at T0 and T2 (see 2.6 below).

Patients were allocated in a 1:1:1 ratio to one of the three treatment groups (DLPFC, preSMA or vertex) using a stratified variable block randomization model in Castor Electronic Data Capture (Castor EDC, https://www.castoredc.com/). Participants were stratified based on medication status (current use of SRI). Randomization was carried out following baseline MRI and clinical assessments by a researcher not involved in clinical assessments.

### 2.3 Interventions

#### 2.3.1 rTMS intervention and blinding

After randomization patients received either: 10Hz rTMS at 110% resting motor threshold (RMT) to the left DLPFC; 10Hz rTMS at 110% RMT to the left preSMA; or 10Hz rTMS at 60% RMT to the vertex. All participants received 3000 pulses in 30x10s trains with 30 seconds intertrain intervals(4).

Stimulation location coordinates (Figure 1) for the DLPFC and preSMA groups were individually determined based on tb-fMRI activation during the Tower of London (TOL(5)) task for the DLPFC condition, and the Stop Signal Task (SST(16)) for the preSMA (see section 2.6 and Supplementary Methods for full details of tasks and contrasts). All participants in the vertex group were stimulated at MNI coordinates [0, -34, 72](4). Neuronavigation to the individualized stimulation location at every rTMS treatment session was carried out using the Localite neuronavigation system (Localite GmbH, Bonn, Germany).

**Figure 1:**
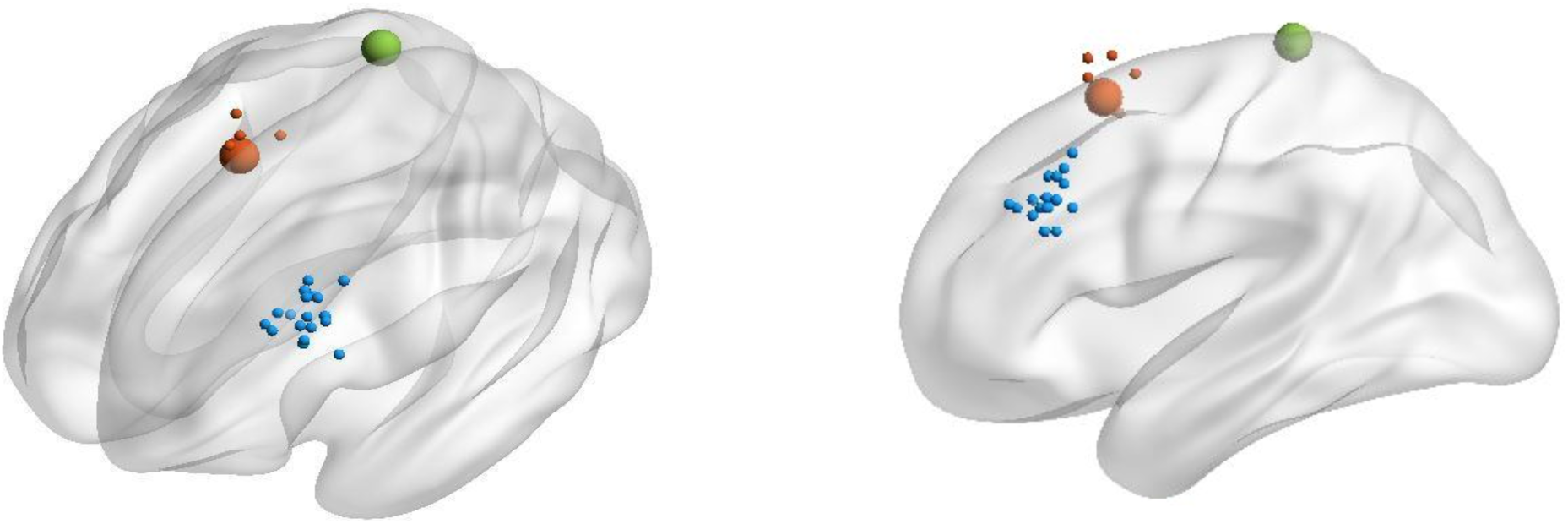
rTMS stimulation locations. Dorsolateral prefrontal cortex group (n=19) in blue (average MNI coordinate -50, 28, 33); pre-supplementary motor area group (n=23) in red (large sphere indicates literature-derived stimulation coordinate (-4, 14, 58), where 74% of the group received stimulation); and the vertex group (n=19) coordinate in green (0, -34, 72).

Participants, psychotherapists and clinical assessors were blinded to rTMS condition; rTMS therapists were unblinded but were instructed not to discuss the treatment conditions with participants or psychotherapists and did not take part in clinical assessments or ERP sessions.

#### 2.3.2 Exposure and response prevention (ERP)

ERP sessions started within 10 minutes after the rTMS session was completed; each session lasted 60 minutes. Psychotherapists followed a protocolized treatment manual, including two introductory sessions, 16 exposure sessions (twice per week, directly following rTMS), one evaluation session, and one relapse prevention session, within ∼10 weeks.

### 2.4 Sample size and outcome measures

Our sample consisted of 61 participants who completed the treatment with clinical outcome measures; 54 of these completed the treatment with both clinical and fMRI outcome measures. See Supplementary Methods for details of the sample size calculation.

Our primary outcome measure was change in tb-fMRI activation following DLPFC/preSMA rTMS compared to vertex rTMS, in task-related regions specific to the stimulated brain circuits (i.e., planning-related activation during the TOL for the DLPFC group; inhibition-related activation during the SST for the preSMA group), using a Bayesian region-of-interest (ROI) approach(17).

Our secondary outcome measure was comparison of T0-T2 change in YBOCS score and number of responders (defined as a reduction of ≥35% in YBOCS score(18)) at T2 in preSMA/DLPFC rTMS versus vertex rTMS groups.

Exploratory outcomes included T0-T2 change in TOL and SST performance, T0-T2 change in depression severity, T0-T2 change in TOL taskload and SST error processing ROI-based fMRI activation, T0-T2 change in whole-brain task-related activation, and the association between T0-T2 change in symptom severity and change in ROI-based fMRI activation.

### 2.5 Assessments

OCD symptom severity was assessed using the YBOCS at T0, T1, T2, and T3. ERP homework and in-session ERP exercise adherence was assessed by psychotherapists at every treatment session using the Patient exposure and response prevention Adherence Scale (PEAS)(19). Severity of depressive symptoms was assessed at T0 and T2 using the Beck Depression Index (BDI)(20). The frequency of common rTMS side effects was assessed at T2 with an in-house questionnaire.

### 2.6 Image acquisition and processing

See Supplementary Methods for full details of image acquisition and tasks. Participants underwent tb-fMRI at T0 and T2, during which they carried out the TOL, a planning task in which planning trials of varying task loads are presented interspersed with counting trials; and the SST, a response inhibition task that requires the participant to respond to left and right arrows by pressing left and right response buttons respectively (go trials), or to refrain from pressing the response button when presented with a cross (stop trials). Tb-fMRI data were pre-processed using fMRIprep v21.0.1, an open-source preprocessing application (https://fmriprep.org). Additional preprocessing steps (8 mm smoothing, high-pass filtering, motion regression) were carried out using SPM12 (Wellcome Trust Centre for Neuroimaging, London) prior to first and second-level analysis. Volumes with >0.5mm frame to frame motion were regressed out (‘scannulled’).

### 2.7 First-level functional imaging analyses & regions of interest

First-level analysis consisted of an event-related design for both tasks, with the following contrasts: TOL, planning (all planning conditions>counting) and taskload (TOL difficulty conditions 5>4>3>2>1, to demonstrate activation during increased planning difficulty); SST, response inhibition (successful stop>go) and error processing (unsuccessful stop>successful stop). ROIs were defined by creating spheres around MNI coordinates, derived from recent meta-analyses of activation during the TOL(21) and SST tasks(22,23) (See Tables S2 and S3 for full list of ROIs and coordinates).

### 2.8 Statistical analysis

The analysis plan was pre-registered at https://osf.io/8r2ys. The primary imaging analysis, an ROI-based analysis, was performed by extracting first-level contrast estimates using MarsBar (v0.44(24)) and conducting the statistical analysis with the AFNI toolbox Region-Based Analysis Program through Bayesian Multilevel Modeling (v1.0.10)(17). We carried out Bayesian multilevel (BML) modeling rather than traditional univariate/null hypothesis testing, as this method has better sensitivity for smaller anatomical areas and grouping of data across subjects and ROIs. It also takes account of the non-independence within a network of ROIs chosen based on a common hypothesis, such as in the present study(17). In this approach, data from multiple ROIs are included in a single multilevel model to evaluate effects of interest. We used BML to compare T0-T2 change in activation between DLPFC and vertex and preSMA and vertex groups, and examine the association between T0-T2 change in activation and T0-T2 change in symptom severity and its difference between groups. We used non-informative Gaussian priors or student’s t-distributed priors if the sample was n<20, and continuous variables were standardized. For all analyses, we added age, sex, and medication status as covariates. For analyses examining the relationship between change in activation and change in YBOCS, we added baseline YBOCS as a covariate. In BML, the probability of the hypothesis given the data is represented as a probability density function called the positive posterior distribution (P+). This is calculated by combining the empirical data with a model and prior expectations. For interpretation, a P+ of <0.2 or >0.8 is classified as weak, <.10 or >.90 as moderate, <.05 or >.95 as strong and <.025 or >.975 as very strong. Exploratory whole-brain second-level analyses were performed in SPM12 using a general linear model with an extent threshold of k=10 and *p*<0.001, uncorrected, with age, sex, and medication status as covariates.

Analysis of the secondary outcome measure (change in YBOCS over time) was carried out using linear mixed models in SPSS (v28; IBM Corp., New York, U.S.), controlling for age and sex (and BDI as post-hoc covariate). Other clinical/cognitive analyses were conducted in R (v4.2.1, Vienna, Austria), using ANOVA/chi-squared/Kruskal-Wallis tests as appropriate. Primary and exploratory fMRI analyses were carried out with all participants who completed ≥80% of the rTMS/ERP sessions (completers) and had fMRI scans available at T0 and T2 (fMRI sample); clinical analyses were carried out with all completers who had clinical assessments available at T0 and T2 (per-protocol sample), and including all participants who started the treatment but completed <80% of sessions (intention to treat sample).

## 3. Results

### 3.1 Sample

Sixty-six patients with OCD with incomplete response to first-line treatments were randomized to ERP combined with either DLPFC, preSMA or vertex rTMS. Of these, 64 started (intention to treat sample) and 61 completed the intervention (per-protocol sample), a dropout rate of 5%. Tb-fMRI data were obtained at both T0 and T2 for 54 participants (fMRI sample) (see Figure 2 for flowchart). Clinical and demographic details are presented in Table 1 for the per-protocol sample and in Table S4 for the fMRI sample. Groups were generally well-matched across demographic and baseline clinical characteristics, including medication status, though the preSMA group had a significantly lower mean age than the other groups (vertex: 40.3 ± 11.3; DLPFC: 40.3 ± 12.4; preSMA 31.5 ± 13.3 (H(2)=9.6, p=0.008)), and the vertex group had a lower BDI score than the other groups (vertex: 11.2 ± 6.86; DLPFC: 22.0 ± 11.3; preSMA 21.6 ± 10.2 (χ^2^(2) =13.535, p=0.001)), The vertex group also had lower rates of comorbid depression (vertex: 0/19; DLPFC: 4/19; preSMA: 8/23 (H(2)=7.999, p=0.018)). Given these between-group differences in BDI, we decided *post-hoc* to add it as a covariate to our analyses of changes in symptom severity.

**Figure 2:**
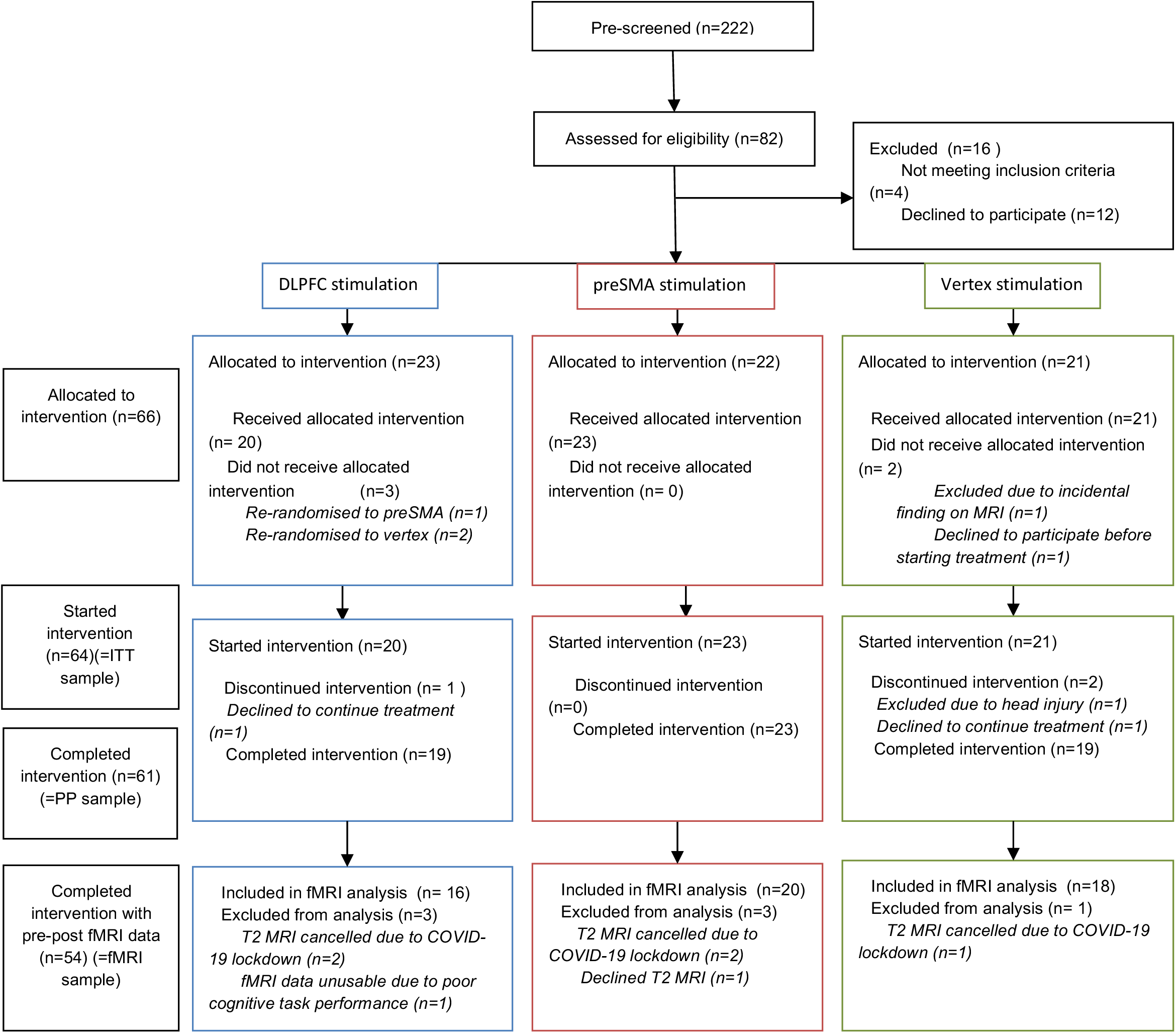
CONSORT diagram: DLPFC = dorsolateral prefrontal cortex; preSMA = pre-supplementary motor area; ITT = intention to treat; PP=per protocol; fMRI = functional magnetic resonance imaging

**Table 1.** demographic and clinical data, per-protocol sample.

|  | All<br>(N=61) | DLPFC<br>(N=19) | preSMA<br>(N=23) | vertex<br>(N=19) | Comparison |
| --- | --- | --- | --- | --- | --- |
| **Demographic data** |  |  |  |  |  |
| **Sex** |  |  |  |  |  |
| male | 22 (36.1%) | 6 (31.6%) | 10 (43.5%) | 6 (31.6%) | $\chi^2(2)=0.880$,<br>p=0.600<sup>a</sup> |
| female | 39 (63.9%) | 13 (68.4%) | 13 (56.5%) | 13 (68.4%) |  |
| **age** |  |  |  |  |  |
| Mean (SD) | 37.0 (12.9) | 40.3 (12.4) | 31.5 (13.3) | 40.3 (11.3) | **H(2)=9.600, p=0.008<sup>b</sup>** |
| **Education<sup>d</sup>** |  |  |  |  |  |
| Median [Min, Max] | 9.00 [4.00, 10.0] | 9.00 [4.00, 10.0] | 9.00 [4.00, 10.0] | 9.00 [6.00, 10.0] | H(2)=1.472, p=0.479<sup>b</sup> |
| **Clinical data** |  |  |  |  |  |
| **Age at symptom onset** |  |  |  |  |  |
| Mean (SD) | 15.4 (7.74) | 15.8 (8.43) | 15.5 (7.30) | 14.9 (7.89) | H(2)=0.00872, p=0.990<sup>b</sup> |
| **Medication status (SRI use)** |  |  |  |  |  |
| unmedicated | 24 (39.3%) | 5 (26.3%) | 9 (39.1%) | 10 (52.6%) | $\chi^2(2)=2.758$,<br>p=0.250<sup>a</sup> |
| medicated | 37 (60.7%) | 14 (73.7%) | 14 (60.9%) | 9 (47.4%) |  |
| **T0 YBOCS** |  |  |  |  |  |
| Mean (SD) | 28.2 (4.58) | 28.8 (5.68) | 29.3 (3.35) | 26.2 (4.24) | F=2.767, p=0.070<sup>c</sup> |
| **T0 BDI** |  |  |  |  |  |
| Mean (SD) | 18.5 (10.7) | 22.0 (11.3) | 21.6 (10.2) | 11.2 (6.86) | **H(2)=13.535, p=0.001<sup>b</sup>** |
| **No. rTMS sessions** |  |  |  |  |  |
| Mean [range] | 16.0 [13.0, 16.0] | 16.0 [13.0, 16.0] | 16.0 [13.0, 16.0] | 16.0 [14.0, 16.0] | H(2)=3.7313, p=0.150<sup>b</sup> |
| **Comorbidities (n, (%))** |  |  |  |  |  |
| Depressive disorder | 12 (19.7) | 4 (21.1) | 8 (34.8) | - | **$\chi^2(2)=7.999$, p=0.018<sup>a</sup>** |
| ADHD | 7 (11.5) | 2 (10.5) | 2 (8.7) | 3 (15.8) | $\chi^2(2)=0.540$, p=0.763<sup>a</sup> |
| General anxiety disorder | 6 (9.8) | 3 (15.8) | 2 (8.7) | 1 (5.3) | $\chi^2(2)=1.241$, p=0.538<sup>a</sup> |
| Panic disorder | 2 (3.3) | 1 (5.3) | - | 1 (5.3) | $\chi^2(2)=1.252$, p=0.535<sup>a</sup> |
| Specific phobia | 2 (3.3) | 2 (10.5) | - | - | $\chi^2(2)=4.571$, p=0.102<sup>a</sup> |
| Alcohol use disorder | 2 (3.3) | 1 (5.3) | 1 (4.3) | - | $\chi^2(2)=0.963$, p=0.618<sup>a</sup> |
| Social anxiety disorder | 1 (1.6) | 1 (5.3) | - | - | $\chi^2(2)=2.247$, p=0.325<sup>a</sup> |
| Body dysmorphic disorder | 1 (1.6) | - | - | 1 (5.3) | $\chi^2(2)=2.247$, p=0.325<sup>a</sup> |
| Anorexia nervosa | 1 (1.6) | 1 (5.3) | - | - | $\chi^2(2)=2.247$, p=0.325<sup>a</sup> |
| PTSD | 1 (1.6) | - | 1 (4.3) | - | $\chi^2(2)=1.680$, p=0.432<sup>a</sup> |

a=chi-squared test; b=kruskal-wallis test; c=one-way anova. ADHD, attention-deficit/hyperactivity disorder; BDI, Beck Depression Index; DLPFC, dorsolateral prefrontal cortex; preSMA, pre-supplementary motor area; PTSD, post-traumatic stress disorder; rTMS, repetitive transcranial magnetic stimulation; SD, standard deviation; SRI, serotonin reuptake inhibitors; T0, baseline; YBOCS, Yale-Brown Obsessive Compulsive Scale

### 3.2 Clinical outcomes

#### 3.2.1 OCD symptom severity

Symptom severity decreased significantly compared to T0 at all three timepoints (T1 mean difference = -7.840, p<0.001, 95% CI [-9.210,-6.470], T2 mean difference = -10.836, p<0.001, 95% CI [-12.504, -9.168]), T3 mean difference = -9.984, p<0.001, 95% CI [-11.832,-8.135] (Figure 3)). There was no statistically significant group by time interaction at any timepoint, both before and after adjusting for age, sex, and baseline BDI (see Tables S5 and S6), indicating no group differences in symptom reduction at any timepoint. Results were similar for the intention to treat sample.

**Figure 3:**
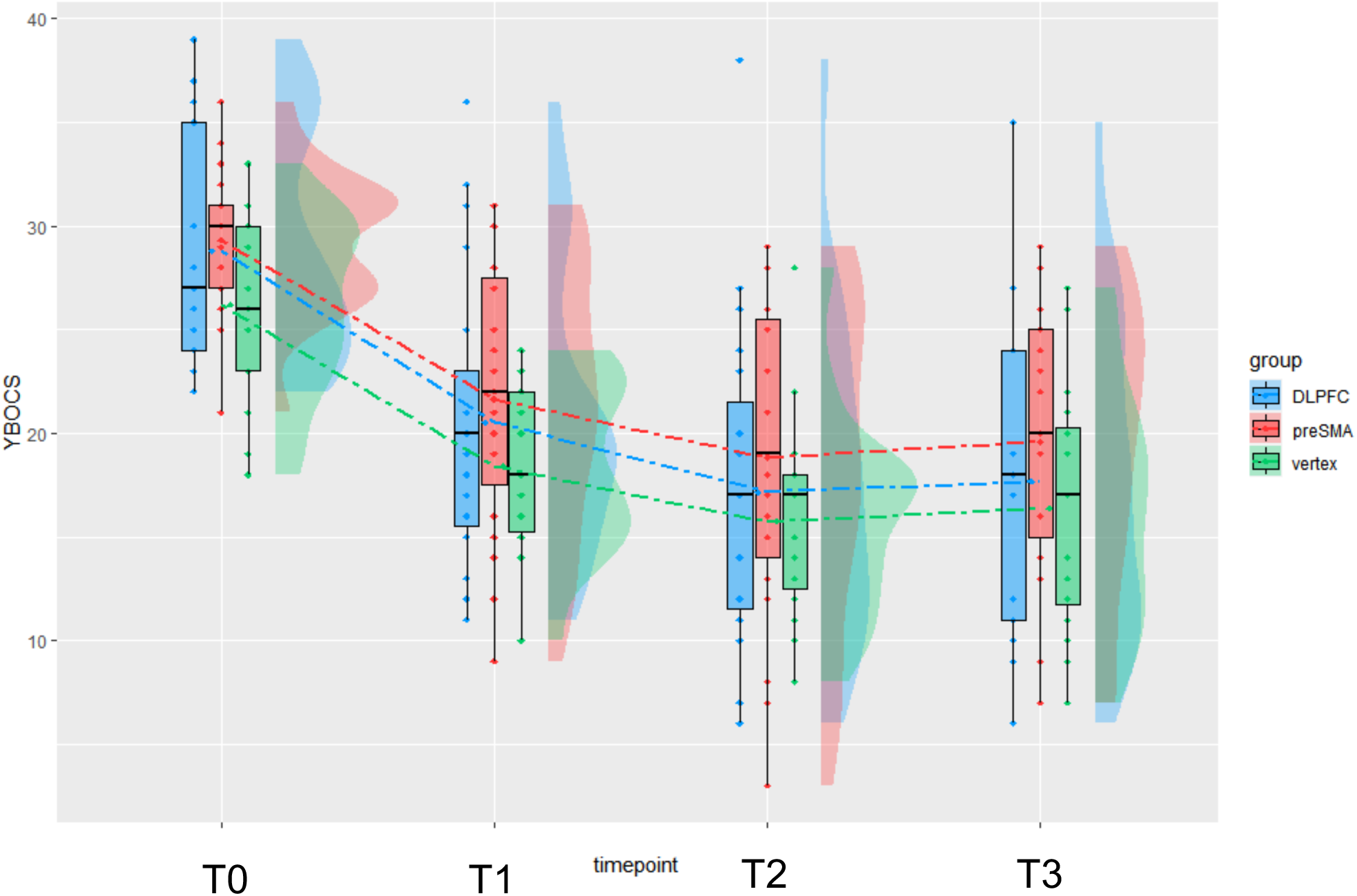
Change in obsessive-compulsive symptom severity over time in all groups: T0=baseline, T1=after 4 weeks of treatment, T2=after 8 weeks of treatment, T3=12 weeks after completing treatment, DLPFC=dorsolateral prefrontal cortex, preSMA = pre-supplementary motor area, YBOCS=Yale-Brown Obsessive Compulsive Scale

#### 3.2.2 Responders/non-responders analysis

Response rate in the entire sample was 57.4% at T2 and 51.9% at T3 (Table 2). Number of responders did not significantly differ between treatment groups for response at T2 (χ^2^(2)=1.377, p=0.502) or T3 (χ^2^(2)=1.125, p=0.570).

**Table 2:** YBOCS scores and response rates across timepoints.

|  | **DLPFC (n=19)** | **preSMA (n=23)** | **Vertex (n=19)** | **All (n=61)** |
| --- | --- | --- | --- | --- |
| **T0 YBOCS**<br>(mean ± SD) | 28.8 ± 5.68 | 29.3 ± 3.35 | 26.2 ± 4.24 | 28.2 ± 4.58 |
| **T1 YBOCS**<br>(mean±SD) | 20.5 ± 7.06 | 21.7 ± 6.57 | 18.4 ± 4.10 | 20.3 ± 6.16 |
| **T2 YBOCS**<br>(mean± SD) | 17.2 ± 7.98 | 18.8 ± 7.80 | 15.7 ± 4.72 | 17.3 ± 7.06 |
| **Responders at**<br>**T2 (n, %)** | 12/19, 63.2% | 11/23, 47.8% | 12/19, 63.1% | 35/61, 57.4% |
| **T3 YBOCS**<br>(mean± SD) | 17.7 ± 7.86 | 19.6 ± 7.13 | 16.4 ± 5.93 | 18.0 ± 7.07 |
| **Responders at**<br>**T3 (n, %)** | 11/19, 57.9% | 8/19 42.1% | 9/16, 56.3% | 28/54, 51.9% |

DLPFC, dorsolateral prefrontal cortex; preSMA, pre-supplementary motor area; SD, standard deviation; T0, baseline; T1, after 4 weeks of treatment; T2, after 8 weeks of treatment; T3, 12 weeks after completing treatment; YBOCS, Yale-Brown Obsessive Compulsive Scale

#### 3.2.3 Other clinical outcomes

Depression symptoms reduced significantly in the entire sample following treatment (T0 BDI 18.5 ± 10.7, T2 BDI 12.2 ± 9.99, W=2492, p=0.0006) and in all three groups separately (Table S7). Average PEAS (ERP adherence) score across all 16 treatment sessions did not differ between the three groups for either homework or in-session exposure exercises (Table S8).

#### 3.2.4 Side effects and safety

At least one side effect (headache, scalp pain, or hearing problems) was reported by 37/60 participants (61.6%), headache being the most frequently reported (32/60, 53.3%). There was no difference between the groups in terms of frequency of the different side effects (Table S9). No serious adverse events were reported.

### 3.3 Task performance

TOL and SST performance data are presented in Table S10. For the TOL, there was no significant effect of group, time, or group by time interaction for either accuracy or reaction time. For the SST, there was a significant decrease in SSRT between T0 and T2 (F(1,52)=4.237, p=.042), but no group effect or group by time interaction.

### 3.4 fMRI outcomes: change in activation following treatment and association with improvement in symptom severity in DLPFC/preSMA vs vertex groups

#### 3.4.1 ROI-based analyses, Bayesian approach

##### 3.4.1.1 TOL planning

There was a specific effect of DLPFC rTMS on planning-related activation: in relation to the vertex group, the DLPFC group showed very strong or strong evidence for a reduction in R inferior parietal cortex (IPC), precuneus, R anterior insula, R DLPFC and L IPC activation after treatment (P+=0.02-0.04) (Table S11; Figure 4). The DLPFC rTMS group, but not the vertex group, also showed strong evidence in the precuneus (P+=0.03) and weak evidence in the R IPC (P+=0.13) for an association between a T0-T2 reduction in activation and symptom improvement; and moderate evidence for an association between a T0-T2 increase in activation and symptom improvement in the R anterior insula (P+=0.94) (Table S11; Figure 5).

**Figure 4:**
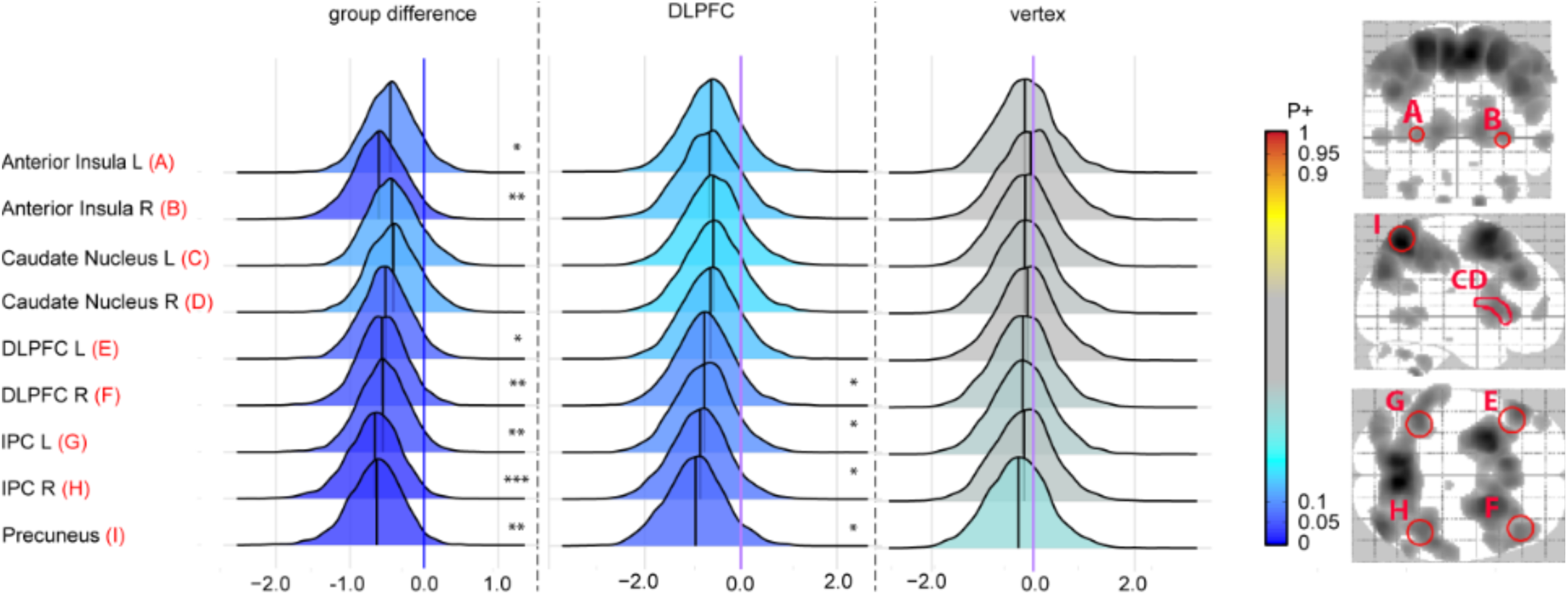
reduction in activation in multiple ROIs during TOL planning contrast following DLPFC rTMS: The right side of the image shows a glass brain (in white) with the darker areas representing brain activation (uncorrected P<0.001, voxel size>5) during planning in all groups combined. The regions of interest are indicated in red with the letters A-I. Posterior distributions show T0-T2 change in activation for DLPFC group (n=16), vertex group (n=18), and group difference (group x time interaction) per ROI. Evidence strength is visualized by the color bar, red indicating increases in activation and blue decreases. For interpretation we added ***= for very strong evidence (P+>0.975 or <0.025), **=strong evidence ((P+>0.95 or <0.05), or *=moderate evidence (P+> 0.90 or <0.10). Corresponding P+ values can be found in Table S11. Abbreviations: DLPFC, dorsolateral prefrontal cortex; IPC, inferior parietal lobe; L, left; PreSMA, presupplementary motor area condition; R, Right

**Figure 5:**
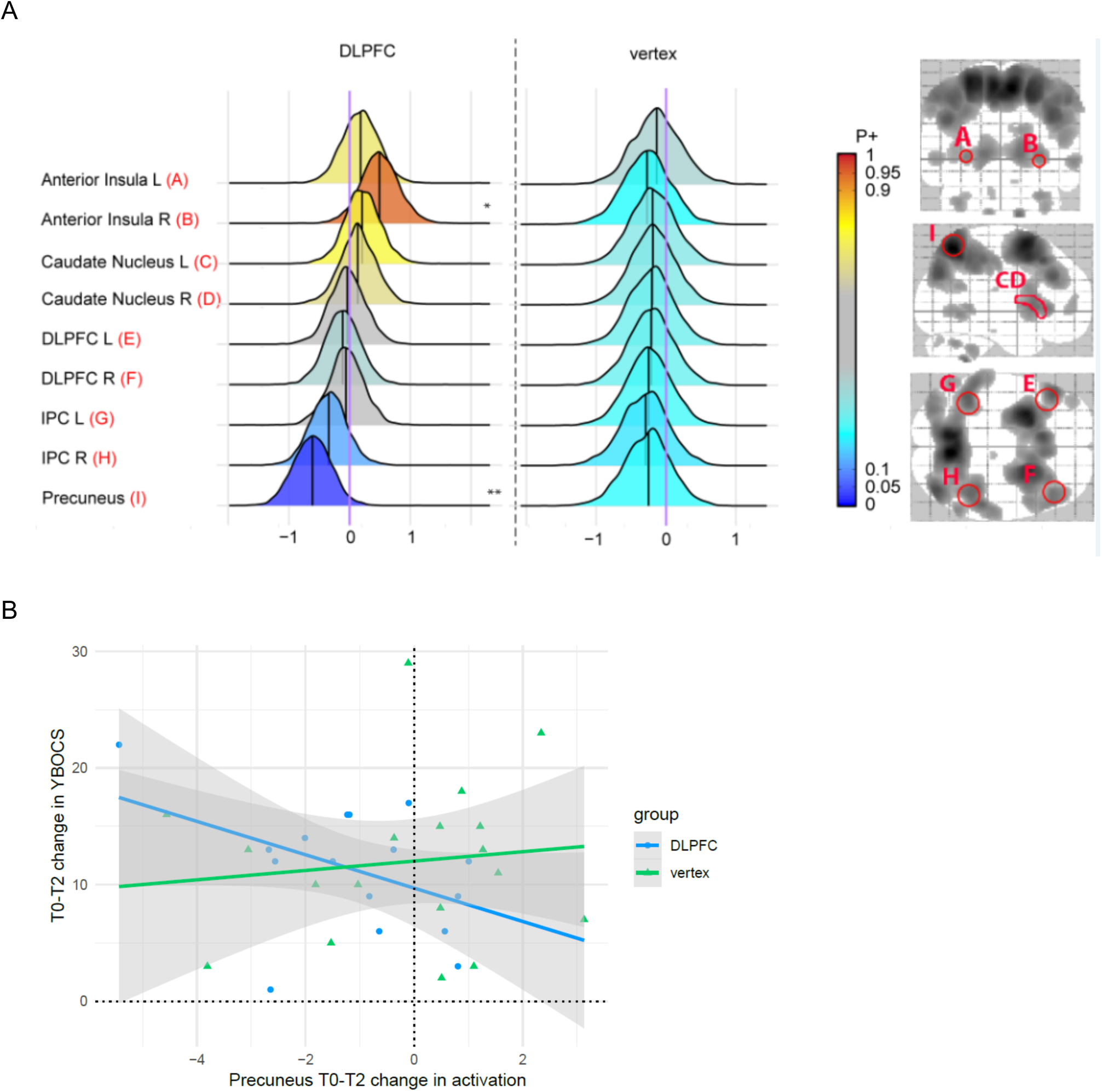
association between T0-T2 change in TOL planning activation and change in YBOCS in DLPFC group vs vertex group: A: Posterior distributions show association between T0-T2 change in YBOCS and change in activation during planning for DLPFC group (n=16), vertex group (n=18), and difference between group associations per ROI. Evidence strength is visualized by the color bar, red indicating positive associations and blue negative. For interpretation we added ***= for very strong evidence (P+>0.975 or <0.025), **=strong evidence ((P+>0.95 or <0.05), or *=moderate evidence (P+> 0.90 or <0.10). Corresponding P+ values can be found in Table S11. B: Scatterplot with visualization of associations in precuneus for DLPFC and vertex groups. Abbreviations: DLPFC, dorsolateral prefrontal cortex; IPC, inferior parietal lobe; L, left; R, Right; YBOCS, Yale-Brown Obsessive-compulsive scale; ROI, region of interest

##### 3.4.1.2 TOL taskload

There was evidence for a differential effect in DLPFC and vertex groups for taskload-related activation. In comparison to the DLPFC group, the vertex group showed an increase in activation across multiple ROIs (very strong evidence for R/L IPC and R precuneus (P+=0.01-0.02); strong evidence for R/L precuneus, (P+=0.03-0.05) and moderate/weak evidence for L anterior insula, BL DLPFC and BL caudate (P+=0.07-0.18) (figure S1). There was, however, little evidence in DLPFC and vertex groups for an association between change in activation and improvement in symptoms (P+=0.44-0.82, Table S12).

##### 3.4.1.3 SST response inhibition

There was no credible evidence for a specific effect on inhibition-related activation in the preSMA group compared with the vertex group (P+=0.51-0.57, table S13). There was, however, moderate to very strong evidence in the vertex group in R anterior insula, R inferior frontal gyrus (IFG), and R parietal cortex (P+=0.90-1.00) for an association between increase in inhibition-related activation and greater symptom reduction, but not in the preSMA group (P+=0.74-0.80, figure 6).

**Figure 6:**
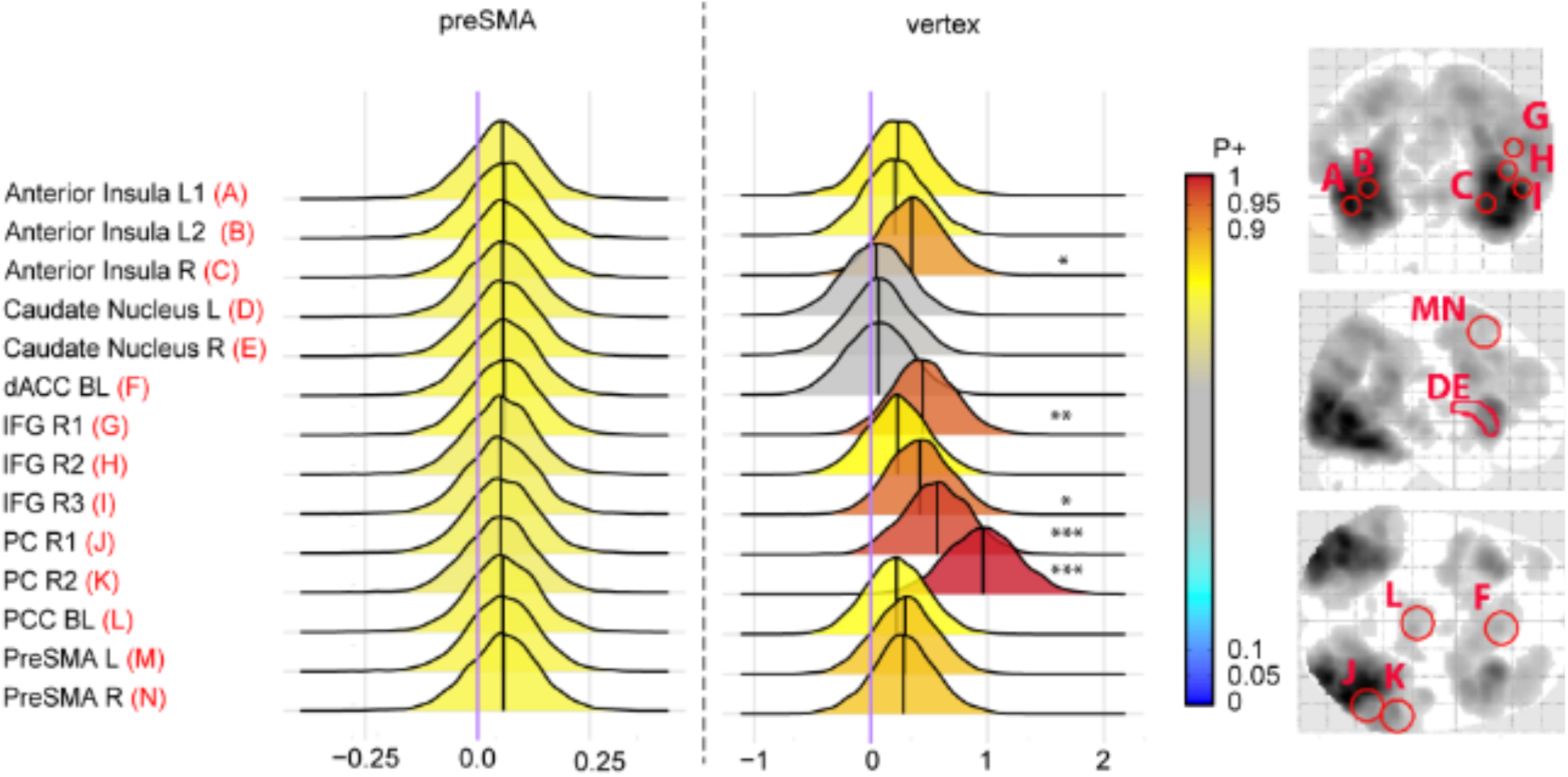
association between T0-T2 change in SST response inhibition activation and change in YBOCS in preSMA group vs vertex group: Posterior distributions show association between T0-T2 change in YBOCS and change in activation during response inhibition for preSMA group (n=20), vertex group (n=18), and difference between group associations per ROI. Evidence strength is visualized by the color bar, red indicating positive associations and blue negative. For interpretation we added ***= for very strong evidence (P+>0.975 or <0.025), **=strong evidence ((P+>0.95 or <0.05), or *=moderate evidence (P+> 0.90 or <0.10). Corresponding P+ values can be found in Table S13. Abbreviations: dACC, dorsal anterior cingulate cortex; DLPFC, dorsolateral prefrontal cortex; IFG, inferior frontal gyrus; L, left; PreSMA, presupplementary motor area; PC, parietal cortex; PCC, posterior cingulate cortex; R, Right, YBOCS, Yale-Brown Obsessive-compulsive scale; ROI, region of interest

##### 3.4.1.4 SST error processing

There was no credible evidence for a specific T0-T2 difference in brain activation in the preSMA group compared to the vertex group (P+=0.37-0.46, table S14). There was weak evidence for an association between decrease in error-related activation and improvement in YBOCS in dACC (P+=0.16) and L anterior insula (P+=0.18) in the preSMA group, but not in the vertex group (P+=0.39, figure 7)

**Figure 7:**
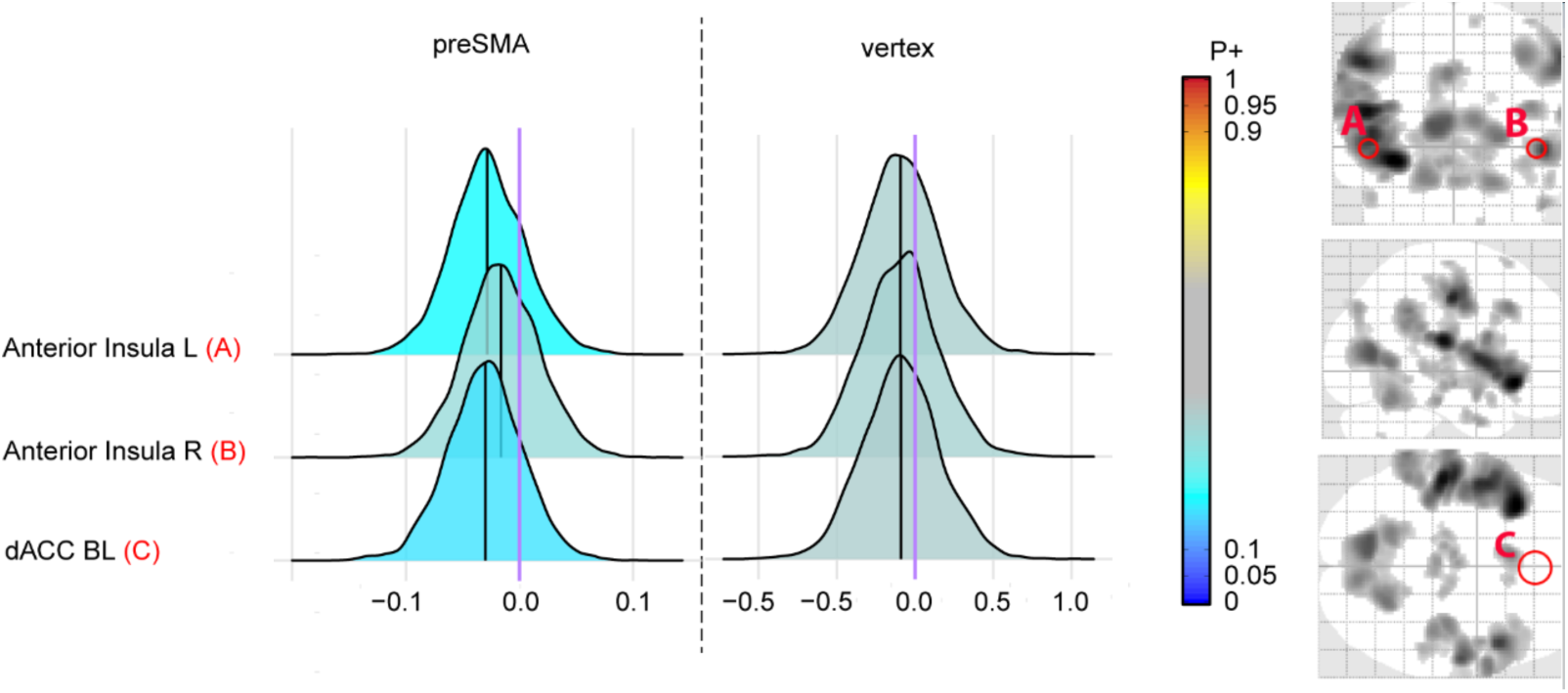
association between T0-T2 change in SST error processing activation and change in YBOCS in preSMA group vs vertex group: Posterior distributions show association between T0-T2 change in YBOCS and change in activation during error processing for preSMA group (n=20), vertex group (n=18), and difference between group associations per ROI. Evidence strength is visualized by the color bar, red indicating positive associations and blue negative. Corresponding P+ values can be found in Table S13. Abbreviations: dACC, dorsal anterior cingulate cortex; L, left; PreSMA, presupplementary motor area; R, Right, YBOCS, Yale-Brown Obsessive-compulsive scale; ROI, region of interest

#### 3.4.2 Whole-brain analyses, GLM approach

Results of the whole-brain analysis were largely consistent with the Bayesian ROI analyses. During the TOL planning and taskload contrasts, the DLPFC group showed a larger decrease in activation than the vertex group in several brain regions (fig S2-S3). Likewise, a relative decrease in activation was observed in the preSMA group relative to the vertex group in the SST response inhibition and error processing contrasts (fig S4-S5). See Supplement for full details.

### 3.5 Change in task-based activation and association with improvement in symptom severity in entire sample (ROI-based analyses, Bayesian approach)

In order to evaluate whether a general treatment effect was present, we repeated the above analyses in all participants together. These are reported fully in the Supplement. While there was little evidence for a T0-T2 change in activation in any contrast, we did find an association between greater symptom improvement and reduced activation following treatment for TOL planning, TOL taskload, and SST error processing, and an association between increased SST inhibition-related activation and greater symptom improvement.

## 4. Discussion

In this proof-of-concept randomized trial, combined rTMS-ERP led to a substantial reduction in symptoms (57.4% responders), but there was no difference in symptom improvement between OCD patients receiving HF rTMS to L DLPFC, L preSMA or vertex. OCD patients showed changes in task-related brain activation following treatment that were specific to rTMS stimulation location and were related to symptom improvement. In the group that received L DLPFC rTMS, we observed a reduction in planning-related activation following rTMS that was associated with a reduction in symptom severity. While we saw no preSMA rTMS-specific change in inhibition- or error-related activation on the group level, we showed that an increase in inhibition-related activation following treatment was related to symptomatic improvement in the vertex group, and a decrease in error-related activation was related to symptomatic improvement in the preSMA-stimulated group. Our whole-brain GLM results largely confirm the results of our Bayesian ROI analysis, indicating decreases in activation during cognitive control in the DLPFC and preSMA groups compared to the vertex group.

The decrease in activation in planning-related areas that was also associated with symptom improvement in the DLPFC group, suggests that HF DLPFC rTMS may result in increased efficiency or reduced effort during planning processes which could underlie response to treatment. This decrease in activation is contrary to our hypothesis, and to the classical assumption that HF rTMS leads to increased excitability of the underlying brain tissue. However, this assumption is largely based on single-session studies in the motor cortex; it is possible that this direction of effect does not necessarily apply outside the motor cortex, or for multiple sessions of rTMS(25). TMS-induced decreases in task-related activation have been demonstrated in previous HF rTMS studies in psychiatric populations(11,26). Multiple sessions of rTMS to the dorsomedial PFC(27) and preSMA(28) in OCD also resulted in reduced resting-state functional connectivity in the targeted networks, suggesting an increase in efficiency. Neurobiological changes following multi-session rTMS may therefore reflect more complex, long-term brain plasticity processes than those observed following single-session rTMS.

We found no group-level changes in inhibition and error-related activation following treatment in any group, similar to the results of Thorsen et al.(29), who also found no group-level change in activation during inhibition or error processing following an intensive ERP treatment for OCD. On the individual level, however, we found a relationship between greater symptom improvement and increased inhibition-related activation after treatment in the vertex group, and a relationship between greater symptom improvement and decreased reactivity to errors in the preSMA group, suggesting differential mechanisms of treatment effect in vertex rTMS + ERP and preSMA rTMS + ERP treatment.

There are several possible explanations for the lack of difference in clinical outcomes between the DLPFC/preSMA and vertex rTMS groups. Firstly, our study was powered to detect tb-fMRI-derived effects (our primary outcome), not clinical effects. All groups also underwent intensive ERP at an expert center for OCD. Additionally, patients may have had high expectations of the therapeutic potential of rTMS, which could have increased the success of the ERP. Secondly, despite randomisation there were some key baseline differences between the groups, notably lower depression scores in the vertex group. Depression has been linked to poorer rTMS and ERP treatment outcomes in OCD(30,31). While correcting for baseline depression scores did not reveal any difference in symptom improvement between groups, there may have still been residual effects that affected the possible degree of improvement in the DLPFC/preSMA rTMS groups. Thirdly, subthreshold rTMS has been documented to have measurable physiological effects on the brain(32,33). There is a possibility that, despite the subthreshold stimulation intensity and the greater distance between the coil and the brain in the vertex condition, some stimulation of the parietal cortex occurred, contributing to a therapeutic effect.

Limitations of this study include insufficient power to detect differences in clinical effects between groups, hindering firm clinical conclusions based on this work. We were also restricted by the number of treatment sessions and our 12-week follow-up period. More treatment sessions may have resulted in a larger difference in clinical outcomes between the groups, and a longer follow-up period may have revealed differences in the persistence of the treatment effect. While our choice of stimulation protocols (HF DLPFC rTMS, HF preSMA rTMS) was based on our previous tb-fMRI studies on cognitive control(4,7), the most frequently used stimulation protocol for the preSMA is LF rTMS, which also has the strongest evidence of clinical effect in OCD(8). This may explain a lack of added symptom improvement, but also limits generalisability and comparability with other rTMS studies in OCD. Our study also has a number of strengths. Firstly, this is the first study that examines tb-fMRI changes following multiple sessions of rTMS for OCD, and is one of only a few studies that use tb-fMRI to describe rTMS treatment effects in psychiatry, possibly offering a more functionally relevant description of treatment effects than resting-state fMRI. Secondly, we included three different rTMS treatment conditions, allowing clinical and neurobiological comparison of multiple rTMS stimulation locations. Thirdly, this trial is the first to combine rTMS with intensive ERP treatment for OCD - our low dropout rate (5%) indicates that this treatment is well tolerated by this population. Finally, we used Bayesian statistics for our primary analysis, which may be more sensitive for our choice of a network of task-relevant ROIs than traditional GLM analyses(17).

In conclusion, this study demonstrates that in OCD, both DLPFC and preSMA HF rTMS in combination with ERP lead to specific decreases in brain activation in targeted cognitive control networks that are associated with symptom improvement, though we found no differences in clinical improvement between preSMA, DLPFC, and vertex rTMS. This finding gives further insights into the complex working mechanisms of rTMS in OCD, and suggests the possibility of inducing specific neurophysiological changes in order to enhance symptom improvement following rTMS. Future studies of rTMS in OCD should 1) continue to add neurobiological outcome measures, especially to multi-session rTMS studies, in order to better characterize the brain plasticity mechanisms underlying the effects of rTMS treatment; 2) perform rTMS trials in combination with ERP with adequate statistical power and sham conditions in order to evaluate the added clinical value of rTMS over and above ERP and to compare different stimulation locations; 3) follow up patients over a longer time period in order to gain knowledge about the long-term clinical and neurobiological effects and cost-effectiveness of this non-invasive brain stimulation technique.

## Data Availability

Datasets are not available to share

## Acknowledgements

Many thanks to all the research interns (Juweiria Abubakar, Shilpa Anand, Julia Biesbroeck, Karolina Brzozowska, Wouter Christiaansen, Farah el Hakkouni, Jorina Holtrop, Eva Leerling, Tinka Louter, Dirk van Paassen, Lieke Pauli, Marjan Ploegaert, Sigrid Roks, Bernard de Roosz, Kirsten Rupert, Nandini P. Sekar, Lars Schlenker, Kim Supit, Ralph Wientjens, Hidde Woerdman, Dennis Zadelhoff, Mimi Elzinga), research assistants (Veerle Daanen, Lara Holzer, Sophie Schubert, Kim Veenman, Rianne Werner, Loïs van ‘t Wout) and therapists (Navin Baitalie, Machteld Blanken, Marlyn Blokland, Kasia Cieslak, Pascale Emmen, Marleen Gideonse, Rosa Maria van den Heuvel, Len Hillen, Sophie Jonker-Teunissen, Kari Jung, Erik van Kemenade, Tokie Kemp, Elsbeth van der Linden, Juliette van der Linden, Isabella Matthews, Floor van der Meer, Larissa Mous, Sterre Rechtuijt, Judith van der Riet, Nina Roosenschoon, Inke Schaap, Pernilla Scheelings, Tomas Sillekens, Joanne van Slooten, Lidewij Smeele, Sabine Stellingwerf, Duygu Talan, Sander Verfaillie, Karin de Vries, Margot van der Wart, Andrea Weeda, Wietske van Wiechen, Elsbeth Zuiker), who helped with data collection and treatments during this study. We thank A. Schweigmann, P. Pouwels and J.P.A Kuijer for MRI technical support and the Patient Association (ADF-stichting) for their valuable contribution. This study was supported by a VIDI grant (91717306, awarded to OvdH) from the Netherlands Organization for Health Research (NWO/ZonMw).

SMDDF was financially supported by the Netherlands Organization for Health Research (NWO/ZonMw, VIDI grant (91717306) to OAvdH); TSP is supported by National Health Care Institute (ZINL/ZonMw VeZo grant 80-86200-98-20006); YDvdW by a NIA Award No. 1R01AG058854-01A1, NINDS award 1RO1NS107513-01A1, a Proof of Concept fund from Amsterdam UMC, in addition to the NWO/ZonMw VIDI grant (91717306), and ZINL/ZonMw VeZo grant (80-86200-98-20006), OAvdH is supported by the National Institute of Mental Health (NIMH) R01 grant (R01MH113250-01) and International OCD Foundation (IOCDF) Innovation grant.

## Disclosures

The authors report no biomedical financial interests or potential conflicts of interest.

This work has been previously published on the following preprint server and presented at the Society of Biological Psychiatry meeting in April 2023

