## Supplementary material for "TMS-induced plasticity improving cognitive control in OCD I: Clinical and neuroimaging outcomes from a randomised trial of rTMS for OCD"

Supplementary information

Supplementary methods:

1.1 Further details of motor threshold determination, rTMS treatment and ERP treatment

The resting motor threshold (RMT) of the left motor cortex was determined once, within 2 weeks prior to starting rTMS treatment. Single TMS pulses were applied, gradually increasing in intensity, until five out of 10 pulses resulted in a right index finger motor-evoked potential of at least 50µV as measured using electromyography of the first dorsal interosseus muscle.

RTMS/MRI exclusion criteria were as follows: pregnancy, ferromagnetic metal implants, implanted electrical devices, personal/family history of epilepsy, other neurological disorders or brain lesions, use of proconvulsive medications, severe heart disease, comorbid substance use or dependence. rTMS treatments were administered using a Magstim Rapid2 TMS stimulator (Magstim, Whitland, Wales, UK) with a Magstim Double 70mm air film coil.

ERP treatments: Each patient worked with two different therapists during the treatment, and therapists received monthly supervision by an expert in CBT for OCD (PvO). Exposure sessions consisted of therapist-aided exposure exercises, and self-guided exposure exercises were assigned as between-session homework.

1.2 Image acquisition and description of fMRI tasks

Participants were scanned on a GE Signa HDxT 3T MRI scanner using a 32-channel head coil (General Electric, Milwaukee, U.S). All functional MRI (fMRI) scans were acquired using a gradient echo-planar imaging sequence (TR = 2.2 s; TE = 26 ms; 64x64 matrix; field of view 21.1cm; flip angle = 80°) with 42 ascending slices per volume (3.3 x 3.3 mm in-plane resolution; slice thickness = 3.0 mm; interslice gap = 0.3 mm). Anatomical MRI was acquired using a 3D T1-weighted structural magnetization-prepared rapid acquisition gradient-echo (MPRAGE; TR = 6.9 ms; TI = 900 ms; TE = 3.0 ms; 256x256 matrix; 1 mm3 isotropic resolution; 168 sections)

All participants carried out the Tower of London (TOL) task and Stop-signal Task (SST) during functional MRI (fMRI). In both cases, task stimuli were presented and responses were recorded using E-prime software (Psychological Software Tools, Pittsburgh, PA) and an MRI compatible button box. The TOL and SST were practiced before the start of the scan session to reduce learning effects.

*Tower of London task*

This task is described in full detail elsewhere(1). Briefly, five different planning conditions, ranging from one to five moves, and one baseline counting condition were presented in a pseudorandomized order (in which each trial of 3 or more moves was followed by a baseline trial). Participants were prompted to either count the number of moves required to move from a starting situation to a goal situation of beads on sticks in a certain configuration (planning conditions) or count the number of beads on two sets of three sticks (counting condition). They had two possible choices of answer which they selected using left or right response buttons. Stimulus presentation was self-paced, with a maximum response time of 60 seconds, and the entire task length was capped at 15 minutes. Participants with an accuracy of <50% on 2 or more conditions were excluded from the imaging analyses.

*Stop-Signal Task*

This task is described in full detail elsewhere(2). Briefly, participants responded to the direction of an arrow pointing left or right by pressing the left or right response buttons (Go trials). 25% of trials were stop trials, presented pseudorandomly, in which a cross would be superimposed on the arrow after a variable delay. Participants were instructed to refrain from responding to the direction of the arrow when the cross appeared. The delay duration was continuously adapted by a staircase-tracking mechanism to ensure an approximately 50% inhibition on all stop trials. GO trials had a fixed duration of 1500 ms and the intertrial interval was jittered between 900 and 1300 ms. 250 trials were presented in 13 minutes, 62 of which were stop trials. Because response latencies gradually increased during the task, stop-signal response times (SSRTs) were estimated separately in 4 smaller blocks (each block consisting of at least 50 trials) and subsequently averaged(3,4). Participants with a go-trial error percentage >40% or stop-trial errors <25% were excluded.

1.3 Image processing for definition of rTMS stimulation targets

Functional images were processed using SPM 12 (Wellcome Trust Centre for Neuroimaging, London). Images were manually reoriented to the T1 scan. During preprocessing, the first three volumes were discarded, and slice timing correction, scanulling of >2mm of movement, normalization to MNI space, and spatial smoothing using an 8 mm Gaussian kernel was carried out. To remove low-frequency noise, a high-pass filter (128-second cutoff period) was applied. Only correct trials were included for task analysis. Participants' movement parameters were included in the model as regressors of no interest. Imaging data were analyzed in the context of the general linear model.

1.4 Creation of stimulation targets

For the DLPFC condition, the peak voxel in the left DLPFC (defined as Brodmann areas 9 and 46(5)) during the planning contrast (all planning conditions > baseline) was determined, and the stimulation target placed on the closest gyrus in the participant's T1 anatomical scan. For the preSMA condition, the local maximum of activation in the left preSMA (defined as the medial gyrus of Brodmann area 6 anterior to the anterior commissure(6)) during the response inhibition contrast (successful stop trials > successful go trials) of the SST were used to determine the stimulation target for the preSMA condition. If no suitable local maximum could be located for the DLPFC or preSMA groups at a statistical threshold of $P<0.01$ uncorrected, the following literature coordinates (in MNI space) were used: i. Left DLPFC: -40, 28, 30(7) ii. Left preSMA: -4, 14, 58(8). For all participants in the vertex group the following MNI coordinates were used: 0, -34, 72(9). Once the individualized stimulation coordinate was defined, a 5mm ROI was created, warped from MNI to subject space and overlaid on a T1 MRI scan of the individual participant, allowing navigation to the individualized stimulation location at every rTMS treatment session using the Localite neuronavigation system (Localite GmbH, Bonn, Germany)

1.5 Sample size

We based our initially planned sample size of 75 (25 per intervention group) on a previous study(9) examining the fMRI effects of a single session of DLPFC rTMS in OCD (n1=19, n2=20, plus 5 participants per group to allow for dropouts). Following lower than expected dropout rates during the study, we re-evaluated our sample size by calculating the minimum detectable effect size (MDES) based on a total sample of 53 (n1=17, n2=18, n3=18), (d=0.928) which was similar to the MDES of de Wit et al 2015(9) (d=0.922), and would still allow us to detect the required effect size. We therefore stopped data collection after reaching an fMRI sample of n=54 (n1=16, n2=20, n3=18).

1.6 Detailed fMRIprep preprocessing boilerplate for T0-T2 activation analysis

Results included in this manuscript come from preprocessing performed using fMRIPrep 21.0.1(10,11); RRID:SCR_016216, which is based on Nipype 1.6.1(12,13); RRID:SCR_002502.

*Preprocessing of B0 inhomogeneity mappings*

A total of 2 fieldmaps were available per subject. A B0-nonuniformity map (or fieldmap) was estimated based on two (or more) echo-planar imaging (EPI) references with topup ((14); FSL 6.0.5.1:57b01774).

*Anatomical data preprocessing*

A total of 2 T1-weighted (T1w) images were found within the input BIDS dataset. All of them were corrected for intensity non-uniformity (INU) with N4BiasFieldCorrection (15), distributed with ANTs 2.3.3 ((16), RRID:SCR_004757). The T1w-reference was then skull-stripped with a Nipype implementation of the antsBrainExtraction.sh workflow (from ANTs), using OASIS30ANTs as target template. Brain tissue segmentation of cerebrospinal fluid (CSF), white-matter (WM) and gray-matter (GM) was performed on the brain-extracted T1w using fast (FSL 6.0.5.1:57b01774,

RRID:SCR_002823, (17)). A T1w-reference map was computed after registration of 2 T1w images (after INU-correction) using mri_robust_template (FreeSurfer 6.0.1, (18)). Brain surfaces were reconstructed using recon-all (FreeSurfer 6.0.1, RRID:SCR_001847,(19)), and the brain mask estimated previously was refined with a custom variation of the method to reconcile ANTs-derived and FreeSurfer-derived segmentations of the cortical gray-matter of Mindboggle (RRID:SCR_002438,(20)). Volume-based spatial normalization to two standard spaces (MNI152NLin6Asym, MNI152NLin2009cAsym) was performed through nonlinear registration with antsRegistration (ANTs 2.3.3), using brain-extracted versions of both T1w reference and the T1w template. The following templates were selected for spatial normalization: FSL's MNI ICBM 152 non-linear 6th Generation Asymmetric Average Brain Stereotaxic Registration Model [(21), RRID:SCR_002823; TemplateFlow ID: MNI152NLin6Asym], ICBM 152 Nonlinear Asymmetrical template version 2009c [(22), RRID:SCR_008796; TemplateFlow ID: MNI152NLin2009cAsym].

*Functional data preprocessing*

For each of the 4 BOLD runs per subject (across all tasks and sessions), the following preprocessing was performed. First, a reference volume and its skull-stripped version were generated using a custom methodology of fMRIPrep. Head-motion parameters with respect to the BOLD reference (transformation matrices, and six corresponding rotation and translation parameters) are estimated before any spatiotemporal filtering using mcflirt (FSL 6.0.5.1:57b01774,(23)). The estimated fieldmap was then aligned with rigid-registration to the target EPI (echo-planar imaging) reference run. The field coefficients were mapped on to the reference EPI using the transform. BOLD runs were slice-time corrected to 1.07s (0.5 of slice acquisition range 0s-2.15s) using 3dTshift from AFNI ((24), RRID:SCR_005927). The BOLD reference was then co-registered to the T1w reference using bbregister (FreeSurfer) which implements boundary-based registration(25). Co-registration was configured with six degrees of freedom. Several confounding time-series were calculated based on the preprocessed BOLD: framewise displacement (FD), DVARS and three region-wise global signals. FD was computed using two formulations following Power (absolute sum of relative motions,(26)) and Jenkinson (relative root mean square displacement between affines,(23)). FD and DVARS are calculated for each functional run, both using their implementations in Nipype (following the definitions by (26)). The three global signals are extracted within the CSF, the WM, and the whole-brain masks. Additionally, a set of physiological regressors were extracted to allow for component-based noise correction (CompCor,(27)). Principal components are estimated after high-pass filtering the preprocessed BOLD time-series (using a discrete cosine filter with 128s cut-off) for the two CompCor variants: temporal (tCompCor) and anatomical (aCompCor). tCompCor components are then calculated from the top 2% variable voxels within the brain mask. For aCompCor, three probabilistic masks (CSF, WM and combined CSF+WM) are generated in anatomical space. The implementation differs from that of Behzadi et al. in that instead of eroding the masks by 2 pixels on BOLD space, the aCompCor masks are subtracted a mask of pixels that likely contain a volume fraction of GM. This mask is obtained by dilating a GM mask extracted from the FreeSurfer's aseg segmentation, and it ensures components are not extracted from voxels containing a minimal fraction of GM. Finally, these masks are resampled into BOLD space and binarized by thresholding at 0.99 (as in the original implementation). Components are also calculated separately within the WM and CSF masks. For each CompCor decomposition, the k components with the largest singular values are retained, such that the retained components' time series are sufficient to explain 50 percent of variance across the nuisance mask (CSF, WM, combined, or temporal). The remaining components are dropped from consideration. The head-motion estimates calculated in the correction step were also placed within the corresponding confounds file. The confound time series derived from head motion estimates and global signals were expanded with the inclusion of temporal derivatives and quadratic terms for each (28). Frames that exceeded a threshold of 0.5 mm FD or 1.5 standardised DVARS were annotated as motion outliers. The BOLD time-series were resampled into standard space, generating a preprocessed BOLD run in MNI152NLin6Asym space. First, a reference volume and its skull-stripped version were generated using a custom methodology of fMRIPrep. The BOLD time-series were resampled onto the following surfaces (FreeSurfer reconstruction nomenclature): fsnative, fsaverage5. Automatic removal of motion artifacts using independent component analysis (ICA-AROMA, (29)) was performed on the preprocessed BOLD on MNI space time-series after removal of non-steady state volumes and spatial smoothing with an isotropic, Gaussian kernel of 6mm FWHM (full-width half-maximum). Corresponding “non-aggresively” denoised runs were produced after such smoothing. Additionally, the “aggressive” noise-regressors were collected and placed in the corresponding confounds file. All resamplings can be performed with a single interpolation step by composing all the pertinent transformations (i.e. head-motion transform matrices, susceptibility distortion correction when available, and co-registrations to anatomical and output spaces). Gridded (volumetric) resamplings were performed using antsApplyTransforms (ANTs), configured with Lanczos interpolation to minimize the smoothing effects of other kernels (30). Non-gridded (surface) resamplings were performed using mri_vol2surf (FreeSurfer).

Many internal operations of fMRIPrep use Nilearn 0.8.1 ((31), RRID:SCR_001362), mostly within the functional processing workflow. For more details of the pipeline, see the section corresponding to workflows in fMRIPrep’s documentation.

Copyright Waiver

The above boilerplate text was automatically generated by fMRIPrep with the express intention that users should copy and paste this text into their manuscripts unchanged. It is released under the CC0 license.

1.7 Background information on BML

For Bayesian Multilevel Modelling (BML), the probability of the hypothesis is estimated given the acquired data, generating a posterior distribution, which can be summarized by a P+ value. Results generated with the Bayesian approach can be interpreted in a natural way by using posterior distributions. The further the posterior distribution is shifted from 0, the stronger the evidence for an association or group difference. Conversely, the closer it is to 0, the stronger the evidence for a negative association or group difference in the opposite direction.

### 2. Supplementary Results

#### *2.1 ROI-based analyses, Bayesian approach, all participants combined*

##### *2.1.1 TOL planning*

Across all participants, there was no credible evidence for a T0-T2 change in planning-related activation (P+ values for all ROIs between 0.47-0.6, Table S11). However, there was moderate evidence for an association between reduction in planning-related activation following treatment and symptom improvement in right inferior parietal cortex (IPC) (P+ 0.08) and bilateral precuneus (P+ 0.09) (Figure S6).

##### *2.1.2 TOL taskload*

For all participants together, there was weak evidence for an increase in TOL taskload-related activation across multiple ROIs (Table S12, P+ 0.86-0.9) following treatment. There was evidence for an association between a reduction in activation and greater improvement in YBOCS scores following treatment across multiple ROIs (P+0.02-0.14; Figure S7, Table S12)

##### *2.1.3 SST response inhibition*

In the entire treatment group, there was little evidence for a change in inhibition-related activation following treatment (P+0.26-0.29, Table S13). There was moderate/weak evidence for an association between increase in activation and improvement in OCD symptoms across all tested ROIs (P+ 0.89-0.92). (Table S13)

##### *2.1.4 SST error processing*

There was no credible evidence for a change in activation during error processing following treatment in the entire sample (P+ 0.38-0.81, table S14). There was weak evidence for a negative relationship between change in dACC activation during error processing and change in symptom severity – a greater decrease in error related dACC activation was associated with a greater decrease in YBOCS ( P+ 0.1, table S14).

2. Supplementary Tables

Table S1: CONSORT checklist

| Section/Topic | Item No | Checklist item | Reported on page No |
|---|---|---|---|
| **Title and abstract** | | | |
| | 1a | Identification as a randomized trial in the title | 1 |
| | 1b | Structured summary of trial design, methods, results, and conclusions | 2 |
| **Introduction** | | | |
| Background and objectives | 2a | Scientific background and explanation of rationale | 3-4 |
| | 2b | Specific objectives or hypotheses | 3-4 |
| **Methods** | | | |
| Trial design | 3a | Description of trial design (such as parallel, factorial) including allocation ratio | 5-6 |
| | 3b | Important changes to methods after trial commencement (such as eligibility criteria), with reasons | n/a |
| Participants | 4a | Eligibility criteria for participants | 5 |
| | 4b | Settings and locations where the data were collected | 5 |
| Interventions | 5 | The interventions for each group with sufficient details to allow replication, including how and when they were actually administered | 6 |
| Outcomes | 6a | Completely defined pre-specified primary and secondary outcome measures, including how and when they were assessed | 7 |
| | 6b | Any changes to trial outcomes after the trial commenced, with reasons | n/a |
| Sample size | 7a | How sample size was determined | 7 |

| Section/Topic | Item No | Checklist item | Reported on page No |
|---|---|---|---|
| | 7b | When applicable, explanation of any interim analyses and stopping guidelines | 7 |
| Randomisation: | | | |
| Sequence generation | 8a | Method used to generate the random allocation sequence | 5-6 |
| | 8b | Type of randomisation; details of any restriction (such as blocking and block size) | 5-6 |
| Allocation concealment mechanism | 9 | Mechanism used to implement the random allocation sequence (such as sequentially numbered containers), describing any steps taken to conceal the sequence until interventions were assigned | 5-6 |
| Implementation | 10 | Who generated the random allocation sequence, who enrolled participants, and who assigned participants to interventions | 5-6 |
| Blinding | 11a | If done, who was blinded after assignment to interventions (for example, participants, care providers, those assessing outcomes) and how | 6 |
| | 11b | If relevant, description of the similarity of interventions | n/a |
| Statistical methods | 12a | Statistical methods used to compare groups for primary and secondary outcomes | 8-9 |
| | 12b | Methods for additional analyses, such as subgroup analyses and adjusted analyses | n/a |
| **Results** | | | |
| Participant flow (a diagram is strongly recommended) | 13a | For each group, the numbers of participants who were randomly assigned, received intended treatment, and were analysed for the primary outcome | 10 |
| | 13b | For each group, losses and exclusions after randomisation, together with reasons | 10, 27 |

| Section/Topic | Item No | Checklist item | Reported on page No |
|---|---|---|---|
| Recruitment | 14a | Dates defining the periods of recruitment and follow-up | 5 |
| | 14b | Why the trial ended or was stopped | n/a |
| Baseline data | 15 | A table showing baseline demographic and clinical characteristics for each group | 32 |
| Numbers analysed | 16 | For each group, number of participants (denominator) included in each analysis and whether the analysis was by original assigned groups | 11 |
| Outcomes and estimation | 17a | For each primary and secondary outcome, results for each group, and the estimated effect size and its precision (such as 95% confidence interval) | 11-14 |
| | 17b | For binary outcomes, presentation of both absolute and relative effect sizes is recommended | 11-14 |
| Ancillary analyses | 18 | Results of any other analyses performed, including subgroup analyses and adjusted analyses, distinguishing pre-specified from exploratory | 16 |
| Harms | 19 | All important harms or unintended effects in each group (for specific guidance see CONSORT for harms[28]) | 17 |
| **Discussion** | | | |
| Limitations | 20 | Trial limitations, addressing sources of potential bias, imprecision, and, if relevant, multiplicity of analyses | 17-20 |
| Generalisability | 21 | Generalisability (external validity, applicability) of the trial findings | 17-20 |
| Interpretation | 22 | Interpretation consistent with results, balancing benefits and harms, and considering other relevant evidence | 17-20 |
| **Other information** | | | |

| Section/Topic | Item No | Checklist item | Reported on page No |
|---|---|---|---|
| Registration | 23 | Registration number and name of trial registry | 5 |
| Protocol | 24 | Where the full trial protocol can be accessed, if available | n/a |
| Funding | 25 | Sources of funding and other support (such as supply of drugs), role of funders | 21 |

Table S2 Coordinates used for ROI analyses of the TOL task

| Brain region | MNI coordinates for planning contrast(32) (x,y,z) | MNI coordinates for taskload contrast(32) (x,y,z) | |
|---|---|---|---|
| L DLPFC* | −40.6, 31.8, 30.6 | | −43.2, 32.8, 30.9 |
| R DLPFC* | 41.4, 35.5, 29.9 | | 42.2, 38.7, 27.3 |
| L caudate nucleus** | AAL ROI | | AAL ROI |
| R caudate nucleus** | AAL ROI | | AAL ROI |
| Bilateral Precuneus* | 0.0, −61.5, 57.3 | L | −11.1, −57.7, 60.9 |
| | - | R1 | 8.3, −59.3, 58.4 |
| | - | R2 | 42.4, −74.8, 39.3 |
| L anterior insula | −31.0, 23.8, −0.9 | | −32.9, 23.1, −4.5 |
| R anterior insula | 33.1, 24.3, −4.2 | | - |
| L IPC* | −38.2, −45.6, 44.9 | 1 | −53.8, −40.7, 48.8 |
| | - | 2 | −40.5, −51.9, 51.0 |
| R IPC* | 46.3, −39.8, 47.2 | | 51.2, −42.8, 46.7 |

Abbreviations: AAL, automated anatomical labeling; DLPFC, dorsolateral prefrontal cortex; IPC, inferior parietal cortex; L, Left; MNI, Montreal neurological institute; R, Right; ROI, Region of interest; TOL, Tower of London.

*=10 mm spheres for larger cortical areas; all other spheres had a radius of 5mm

** For the left and right caudate nucleus, we used the automated anatomical labeling (AAL) atlas(33), to avoid overlap with neighboring subcortical regions.

Table S3 Coordinates used for ROI analyses of the SST task

| Brain region | MNI coordinates for response inhibition contrast(34) (x, y, z) | MNI coordinates for error processing contrast(8) (x,y,z) |
|---|---|---|
| L preSMA | -4,16,48⁺ | - |
| R preSMA | 4,16,48 | - |
| L anterior insula 1 | -40, 16, -4 | - 48,16,-2 |
| L anterior insula 2 | -32, 20, 2 | - |
| R anterior insula | 34, 22, -4 | 44,14,2 |
| R IFG 1 | 50, 12, 28 | - |
| R IFG 2 | 48, 16, 16 | - |
| R IFG 3 | 56, 20, 4 | - |
| L caudate nucleus** | AAL ROI | - |
| R caudate nucleus** | AAL ROI | - |
| R parietal cortex 1 | 58, -48, 32 | - |
| R parietal cortex 2 | 62,-42,26 | - |
| bilateral posterior cingulate cortex* | 2, -24, 32 | - |
| bilateral dACC* | 6, 30, 32 | 0,34,32 |

Abbreviations: AAL, automated anatomical labeling; dACC, dorsal anterior cingulate cortex; IFG, inferior frontal gyrus L, Left; MNI, Montreal neurological institute; R, Right

*=10 mm spheres for larger cortical areas; all other spheres had a radius of 5mm

**For the left and right caudate nucleus, we used the automated anatomical labeling (AAL) atlas(33), to avoid overlap with neighboring subcortical regions.

Table S4 – demographic and clinical data, fMRI sample

| | Overall (N=54) | DLPFC (N=16) | preSMA (N=20) | vertex (N=18) | Comparison |
|---|---|---|---|---|---|
| **Demographic data** | | | | | |
| **Sex** | | | | | |
| male | 19 (35.2%) | 5 (31.3%) | 9 (45.0%) | 5 (27.8%) | $\chi^2$=1.387, |
| female | 35 (64.8%) | 11 (68.8%) | 11 (55.0%) | 13 (72.2%) | p=0.500[a] |
| **age** | | | | | |
| Mean (SD) | 36.4 (12.6) | 39.9 (11.3) | 30.0 (12.3) | 40.5 (11.5) | **H(2)=12.608, p=0.002[b]** |
| **education** | | | | | |
| Median [Min, Max] | 9.00 [4.00, 10.0] | 8.50 [4.00, 10.0] | 8.50 [4.00, 10.0] | 9.00 [6.00, 10.0] | H(2)=1.537, p=0.464[b] |
| **Clinical data** | | | | | |
| **Age of symptom onset** | | | | | |
| Mean (SD) | 15.0 (7.77) | 14.6 (8.39) | 15.8 (7.42) | 14.6 (7.98) | H(2)=0.226, p=0.89[b] |
| **Medication status** | | | | | |
| unmedicated | 22 (40.7%) | 5 (31.3%) | 7 (35.0%) | 10 (55.6%) | $\chi^2$=2.506, |
| medicated | 32 (59.3%) | 11 (68.8%) | 13 (65.0%) | 8 (44.4%) | p=0.286[a] |
| **T0 YBOCS** | | | | | |
| Mean (SD) | 28.0 (4.55) | 28.5 (5.49) | 29.3 (3.42) | 26.2 (4.36) | F=2.514, p=0.09[c] |
| **T0 BDI** | | | | | |
| Mean (SD) | 18.2 (11.1) | 22.4 (12.2) | 21.1 (10.8) | 11.4 (7.01) | **H(2)=10.136, p=0.006[b]** |
| **No. rTMS sessions** | | | | | |
| Mean (SD) | 15.7 (0.705) | 15.9 (0.500) | 15.5 (0.946) | 15.9 (0.471) | H(2)=3.991, p=0.136[b] |

| | | | | | |
|---|---|---|---|---|---|
| **Comorbidities (n, (%))** | | | | | |
| Depressive disorder | 10 (18.5) | 3 (18.8) | 7 (35.0) | - | $\chi^2$=7.692, **p=0.021** [a] |
| ADHD | 7 (13.0) | 2 (12.5) | 2 (10.0) | 3 (16.7) | $\chi^2$=0.378, p=0.828 [a] |
| General anxiety disorder | 5 (9.3) | 3 (18.8) | 1 (5.0) | 1 (5.6) | $\chi^2$=2.441, p=0.295 [a] |
| Panic disorder | 2 (3.7) | 1 (6.3) | - | 1 (5.6) | $\chi^2$=1.233, p=0.540[a] |
| Specific phobia | 2 (3.7) | 2 (12.5) | - | - | $\chi^2$=4.933, p=0.085 [a] |
| Alcohol use disorder | 2 (3.7) | 1 (6.3) | 1 (5.0) | - | $\chi^2$=1.077, p=0.584 [a] |
| Social anxiety disorder | 1 (1.9) | 1 (6.3) | - | - | $\chi^2$=2.420, p=0.298 [a] |
| Body dysmorphic disorder | 1 (1.9) | - | - | 1 (5.6) | $\chi^2$=2.038, p=0.361 [a] |
| Anorexia nervosa | 1 (1.9) | 1 (6.3) | - | - | $\chi^2$=2.420, p=0.298 [a] |
| PTSD | 1 (1.9) | - | 1 (5.0) | - | $\chi^2$=1.732, p=0.421 [a] |

a=chi-squared test; b=kruskal-wallis test; c=one-way anova. ADHD, attention-deficit/hyperactivity disorder; BDI, Beck Depression Index; DLPFC, dorsolateral prefrontal cortex; preSMA, pre-supplementary motor area; PTSD, post-traumatic stress disorder; rTMS, repetitive transcranial magnetic stimulation; SD, standard deviation; SRI, serotonin reuptake inhibitors; T0, baseline; YBOCS, Yale-Brown Obsessive Compulsive Scale

Table S5: group by time interaction for change in YBOCS over time: linear mixed models

| | Parameter | Estimate | Std. Error | df | t | Sig. | 95% Confidence Interval Lower Bound | Upper Bound |
|---|---|---|---|---|---|---|---|---|
| Vertex vs DLPFC | T0 vs T1 | .185 | 1.709 | 58.573 | .109 | .914 | -3.234 | 3.605 |
| | T0 vs T2 | -.272 | 2.083 | 58.000 | -.131 | .896 | -4.442 | 3.898 |
| | T0 vs T3 | .120 | 2.224 | 55.664 | .054 | .957 | -4.335 | 4.575 |
| Vertex vs preSMA | T0 vs T1 | .955 | 1.634 | 58.637 | .585 | .561 | -2.314 | 4.225 |
| | T0 vs T2 | 1.058 | 1.991 | 58.000 | .531 | .597 | -2.927 | 5.042 |
| | T0 vs T3 | 2.432 | 2.186 | 57.623 | 1.112 | .271 | -1.945 | 6.808 |
| DLPFC vs preSMA | T0 vs T1 | .770 | 1.626 | 57.895 | .474 | .638 | -2.485 | 4.025 |
| | T0 vs T2 | 1.330 | 1.991 | 58.000 | .668 | .507 | -2.654 | 5.315 |
| | T0 vs T3 | 2.312 | 2.126 | 55.695 | 1.087 | .282 | -1.947 | 6.570 |

DLPFC, dorsolateral prefrontal cortex; preSMA, pre-supplementary motor area; T0, baseline; T1, after 4 weeks of treatment; T2, after 8 weeks of treatment; T3, 12 weeks after completing treatment; YBOCS, Yale-Brown Obsessive Compulsive Scale

Table S6: group by time interaction for change in YBOCS over time while controlling for BDI, age, sex

| | Parameter | Estimate | Std. Error | df | t | Sig. | 95% Confidence Interval Lower Bound | Upper Bound |
|---|---|---|---|---|---|---|---|---|
| Vertex vs DLPFC | T0 vs T1 | -.639 | 1.625 | 58.837 | -.393 | .695 | -3.890 | 2.612 |
| | T0 vs T2 | -1.183 | 1.943 | 58.310 | -.609 | .545 | -5.071 | 2.705 |
| | T0 vs T3 | -1.187 | 2.180 | 56.667 | -.544 | .588 | -5.552 | 3.179 |
| Vertex vs preSMA | T0 vs T1 | .234 | 1.558 | 59.600 | .150 | .881 | -2.884 | 3.351 |
| | T0 vs T2 | .311 | 1.863 | 59.006 | .167 | .868 | -3.417 | 4.040 |
| | T0 vs T3 | 1.248 | 2.151 | 58.489 | .580 | .564 | -3.058 | 5.553 |
| DLPFC vs preSMA | T0 vs T1 | .873 | 1.534 | 56.631 | .569 | .572 | -2.200 | 3.946 |
| | T0 vs T2 | 1.494 | 1.841 | 56.679 | .812 | .420 | -2.193 | 5.182 |
| | T0 vs T3 | 2.434 | 2.072 | 55.805 | 1.175 | .245 | -1.716 | 6.584 |

BDI, Beck Depression Index; DLPFC, dorsolateral prefrontal cortex; preSMA, pre-supplementary motor area; T0, baseline; T1, after 4 weeks of treatment; T2, after 8 weeks of treatment; T3, 12 weeks after completing treatment; YBOCS, Yale-Brown Obsessive Compulsive Scale

Table S7: Change in BDI scores between T0 and T2

| | T0 | T2 | comparison |
|---|---|---|---|
| DLPFC | 22.0 ± 11.3 | 13.9 ± 11.4 | W= 264.5, **p=0.015** |
| preSMA | 21.6 ± 10.2 | 15.1 ± 9.99 | W=344.5, **p=0.038** |
| vertex | 11.2 ± 6.86 | 6.95 ± 6.22 | W=248, **p=0.050** |

BDI, Beck Depression Index; DLPFC, dorsolateral prefrontal cortex; preSMA, pre-supplementary motor area; T0, baseline; T2, after 8 weeks of treatment

Table S8: Average PEAS scores

| | All | DLPFC | preSMA | Vertex | comparison |
|---|---|---|---|---|---|
| average PEAS per session | 6.07 ± 0.777 | 5.97 ± 1.08 | 5.91 ± 0.624 | 6.35 ± 0.485 | $\chi^2(2)=4.53$, $p=0.104$ |
| average PEAS per homework | 5.18 ± 0.963 | 4.99 ± 1.17 | 5.13 ± 0.733 | 5.45 ± 0.978 | $\chi^2(2)=1.68$, $p=0.431$ |

DLPFC, dorsolateral prefrontal cortex; preSMA, pre-supplementary motor area; PEAS, patient exposure/response prevention adherence scale

Table S9: frequency of common rTMS side effects per group

| | All (n=60) | DLPFC (n=19) | preSMA (n=22) | vertex (n=19) | comparison (chi-squared test) |
|---|---|---|---|---|---|
| **Headache** | | | | | |
| none | 28 (45.9%) | 8 (42.1%) | 11 (47.8%) | 9 (47.4%) | $\chi^2(6)=5.598$, p=0.470 |
| light | 14 (23.0%) | 6 (31.6%) | 5 (21.7%) | 3 (15.8%) | |
| moderate | 14 (23.0%) | 5 (26.3%) | 3 (13.0%) | 6 (31.6%) | |
| severe | 4 (6.6%) | 0 (0%) | 3 (13.0%) | 1 (5.3%) | |
| **Scalp pain** | | | | | |
| none | 42 (68.9%) | 12 (63.2%) | 17 (73.9%) | 13 (68.4%) | $\chi^2(4)=1.324$, p=0.857 |
| light | 14 (23.0%) | 5 (26.3%) | 4 (17.4%) | 5 (26.3%) | |
| moderate | 4 (6.6%) | 2 (10.5%) | 1 (4.3%) | 1 (5.3%) | |
| **Hearing problems*** | | | | | |
| none | 52 (85.2%) | 16 (84.2%) | 19 (82.6%) | 17 (89.5%) | $\chi^2(4)=4.729$, p=0.316 |
| light | 5 (8.2%) | 2 (10.5%) | 3 (13.0%) | 0 (0%) | |
| moderate | 3 (4.9%) | 1 (5.3%) | 0 (0%) | 2 (10.5%) | |

* ringing ears or a change in hearing; DLPFC, dorsolateral prefrontal cortex; preSMA, pre-supplementary motor area

Table S10: Performance on TOL and SST at baseline and following treatment

| | T0 | | | | T2 | | | |
|---|---|---|---|---|---|---|---|---|
| | DLPFC (n=16) | preSMA (n=20) | Vertex (n=18) | All (n=54) | DLPFC (n=16) | preSMA (n=20) | Vertex (n=18) | All (n=54) |
| TOL accuracy (%, (sd)) | 88.4 (10.5) | 86.6 (12.4) | 85.3 (11.3) | 86.7 (11.4) | 87.0 (11.2) | 86.0 (11.8) | 85.8 (10.9) | 86.2 (11.1) |
| TOL reaction time (mean s, (sd)) | 9.85 (3.50) | 9.12 (2.60) | 10.7 (2.82) | 9.86 (2.98) | 9.52 (3.80) | 9.11 (4.13) | 9.80 (3.61) | 9.47 (3.80) |
| SSRT (mean s, (sd)) | 213 (49.4) | 212 (63.7) | 210 (71.9) | 212 (61.5) | 184 (80.0) | 190 (58.1) | 185 (52.4) | 186 (62.7) |

DLPFC, dorsolateral prefrontal cortex; preSMA, pre-supplementary motor area; SST, stop-signal task; SSRT, stop-signal response time; T0, baseline; T2, following 8 weeks of treatment; TOL, Tower of London

Table S11: TOL Planning BML results

| | T0-T2 Δactivation | | | | association Δactivation and ΔYBOCS | | |
|---|---|---|---|---|---|---|---|
| **ROI** | **P+ all participants (n=54)** | **P+DLPFC (n=16)** | **P+Vertex (n=18)** | **P+ group by time interaction** | **P+ all participants (n=54)** | **P+DLPFC (n=16)** | **P+Vertex (n=18)** |
| Anterior insula L | 0.562 | 0.144 | 0.398 | 0.098* | 0.313 | 0.725 | 0.364 |
| Anterior insula R | 0.481 | 0.139 | 0.462 | 0.037** | 0.330 | 0.939* | 0.209 |
| Caudate nucleus L | 0.598 | 0.165 | 0.390 | 0.108 | 0.348 | 0.752 | 0.303 |
| Caudate nucleus R | 0.562 | 0.167 | 0.388 | 0.116 | 0.323 | 0.679 | 0.302 |
| DLPFC L | 0.570 | 0.141 | 0.421 | 0.056* | 0.232 | 0.451 | 0.269 |
| DLPFC R | 0.489 | 0.090* | 0.376 | 0.040** | 0.207 | 0.346 | 0.266 |
| IPC L | 0.509 | 0.092* | 0.362 | 0.044** | 0.243 | 0.419 | 0.231 |
| IPC R | 0.471 | 0.074* | 0.381 | 0.021*** | 0.081* | 0.125 | 0.192 |
| Precuneus BL | 0.530 | 0.057* | 0.325 | 0.025** | 0.090* | 0.028** | 0.209 |

Corresponding P+ values of Figures 4, 5 and S1, showing the change in activation (**Δactivation)** during TOL planning between timepoints T0 and T2, and the association between **Δactivation** and change in symptom severity (**ΔYBOCS**). For interpretation we added ***= for very strong evidence (P+>0.975 or <0.025), **=strong evidence ((P+>0.95 or <0.05), or *=moderate evidence (P+> 0.90 or <0.10). Abbreviations: BL, Bilateral;DLPFC, dorsolateral prefrontal cortex; IPC, inferior parietal lobe; L, left; R, Right; ROI, Region of interest; TOL, Tower of London

Table S12 TOL Taskload BML results

| | T0-T2 Δactivation | | | | association Δactivation and ΔYBOCS | | |
|---|---|---|---|---|---|---|---|
| **ROI** | **P+ all participants (n=54)** | **P+DLPFC (n=16)** | **P+Vertex (n=18)** | **P+ group by time interaction** | **P+ all participants (n=54)** | **P+DLPFC (n=16)** | **P+Vertex (n=18)** |
| Anterior insula L | 0.888 | 0.387 | 0.728 | 0.067* | 0.431 | 0.825 | 0.817 |
| Caudate nucleus L | 0.871 | 0.421 | 0.654 | 0.185 | 0.251 | 0.771 | 0.752 |
| Caudate nucleus R | 0.862 | 0.383 | 0.681 | 0.113 | 0.345 | 0.786 | 0.766 |
| DLPFC L | 0.872 | 0.357 | 0.707 | 0.071* | 0.103 | 0.786 | 0.669 |
| DLPFC R | 0.895 | 0.400 | 0.739 | 0.068* | 0.072* | 0.761 | 0.595 |
| IPC L1 | 0.882 | 0.290 | 0.808 | 0.010*** | 0.141 | 0.794 | 0.624 |
| IPC L2 | 0.870 | 0.312 | 0.801 | 0.015*** | 0.021*** | 0.785 | 0.707 |
| IPC R | 0.866 | 0.241 | 0.783 | 0.006*** | 0.024*** | 0.680 | 0.749 |
| Precuneus L | 0.872 | 0.347 | 0.732 | 0.047** | 0.109 | 0.773 | 0.532 |
| Precuneus R1 | 0.873 | 0.354 | 0.767 | 0.035** | 0.026** | 0.745 | 0.441 |
| Precuneus R2 | 0.899 | 0.366 | 0.823 | 0.019*** | 0.095* | 0.817 | 0.668 |

Corresponding P+ values of Figures S2 and S3, showing the change in activation (**Δactivation)** during TOL taskload between timepoints T0 and T2, and the association between **Δactivation** and change in symptom severity (**ΔYBOCS)**. For interpretation we added ***= for very strong evidence (P+>0.975 or <0.025), **=strong evidence ((P+>0.95 or <0.05), or *=moderate evidence (P+> 0.90 or <0.10). Abbreviations: BL, Bilateral;DLPFC, dorsolateral prefrontal cortex; IPC, inferior parietal lobe; L, left; R, Right; TOL, Tower of London

Table S13 SST Response inhibition BML results

| | T0-T2 Δactivation | | | | association Δactivation and ΔYBOCS | | |
|---|---|---|---|---|---|---|---|
| **ROI** | **P+ all participants (n=54)** | **P+preSMA (n=20)** | **P+Vertex (n=18)** | **P+ group by time interaction** | **P+ all participants (n=54)** | **P+preSMA (n=20)** | **P+Vertex (n=18)** |
| Anterior insula L1 | 0.281 | 0.331 | 0.365 | 0.514 | 0.924* | 0.763 | 0.811 |
| Anterior insula L2 | 0.293 | 0.362 | 0.359 | 0.562 | 0.913* | 0.760 | 0.778 |
| Anterior insula R | 0.292 | 0.359 | 0.344 | 0.572 | 0.927* | 0.780 | 0.903* |
| Caudate Nucleus L | 0.287 | 0.351 | 0.360 | 0.543 | 0.907* | 0.763 | 0.566 |
| Caudate Nucleus R | 0.285 | 0.366 | 0.361 | 0.558 | 0.906* | 0.769 | 0.604 |
| dACC BL | 0.280 | 0.356 | 0.360 | 0.542 | 0.929* | 0.789 | 0.591 |
| IFG R1 | 0.278 | 0.339 | 0.356 | 0.535 | 0.942* | 0.799 | 0.950** |
| IFG R2 | 0.270 | 0.337 | 0.356 | 0.534 | 0.904* | 0.755 | 0.801 |
| IFG R3 | 0.288 | 0.362 | 0.362 | 0.558 | 0.907* | 0.775 | 0.943* |
| PC R1 | 0.274 | 0.346 | 0.349 | 0.560 | 0.914* | 0.752 | 0.977*** |
| PC R2 | 0.265 | 0.321 | 0.356 | 0.522 | 0.941* | 0.753 | 1.000*** |
| PCC BL | 0.273 | 0.360 | 0.358 | 0.558 | 0.906* | 0.744 | 0.796 |
| PreSMA L | 0.258 | 0.332 | 0.343 | 0.555 | 0.917* | 0.759 | 0.857 |
| PreSMA R | 0.290 | 0.364 | 0.362 | 0.567 | 0.918* | 0.774 | 0.858 |

Corresponding P+ values of Figure 6, showing the change in activation (**Δactivation)** during SST response inhibition between timepoints T0 and T2, and the association between **Δactivation** and change in symptom severity (**ΔYBOCS)**. For interpretation we added ***= for very strong evidence (P+>0.975 or <0.025), **=strong evidence ((P+>0.95 or <0.05), or *=moderate evidence (P+> 0.90 or <0.10). Abbreviations: dACC, dorsal anterior cingulate cortex; IFG, inferior frontal gyrus; L, left; PreSMA, presupplementary motor area; PC, parietal cortex; PCC, posterior cingulate cortex; R, Right; SST, stop-signal task

Table S14 SST Error processing

| | T0-T2 Δactivation | | | | association Δactivation and ΔYBOCS | | |
|---|---|---|---|---|---|---|---|
| | P+ all participants (n=54) | P+preSMA (n=20) | P+Vertex (n=18) | P+ group by time interaction | P+ all participants (n=54) | P+preSMA (n=20) | P+Vertex (n=18) |
| Anterior insula L | 0.380 | 0.643 | 0.654 | 0.436 | 0.148 | 0.183 | 0.363 |
| Anterior insula R | 0.674 | 0.541 | 0.614 | 0.366 | 0.257 | 0.337 | 0.320 |
| dACC BL | 0.810 | 0.601 | 0.610 | 0.458 | 0.111 | 0.161 | 0.405 |

Corresponding P+ values of Figures 7 and S4, showing the change in activation (**Δactivation)** during SST error processing between timepoints T0 and T2, and the association between **Δactivation** and change in symptom severity (**ΔYBOCS)**. For interpretation we added ***= for very strong evidence (P+>0.975 or <0.025), **=strong evidence ((P+>0.95 or <0.05), or *=moderate evidence (P+> 0.90 or <0.10). Abbreviations: BL, bilateral; dACC, dorsal anterior cingulate cortex; L, left; PreSMA, presupplementary motor area; R, Right;  SST, stop-signal task

3. Supplementary Figures

Figure S1: relative increase in in activation in multiple ROIs in vertex group compared to DLPFC group during TOL taskload contrast following DLPFC rTMS: Posterior distributions show T0-T2 change in activation for DLPFC group (n=16), vertex group (n=18), and difference between groups (group x time interaction) per ROI. Evidence strength is visualized by the color bar, red indicating positive associations and blue negative. For interpretation we added ***= for very strong evidence (P+>0.975 or <0.025), **=strong evidence ((P+>0.95 or <0.05), or *=moderate evidence (P+> 0.90 or <0.10). Corresponding P+ values can be found in Table S12. Abbreviations: DLPFC, dorsolateral prefrontal cortex; IPC, inferior parietal lobe; L, left; R, Right; ROI, region of interest

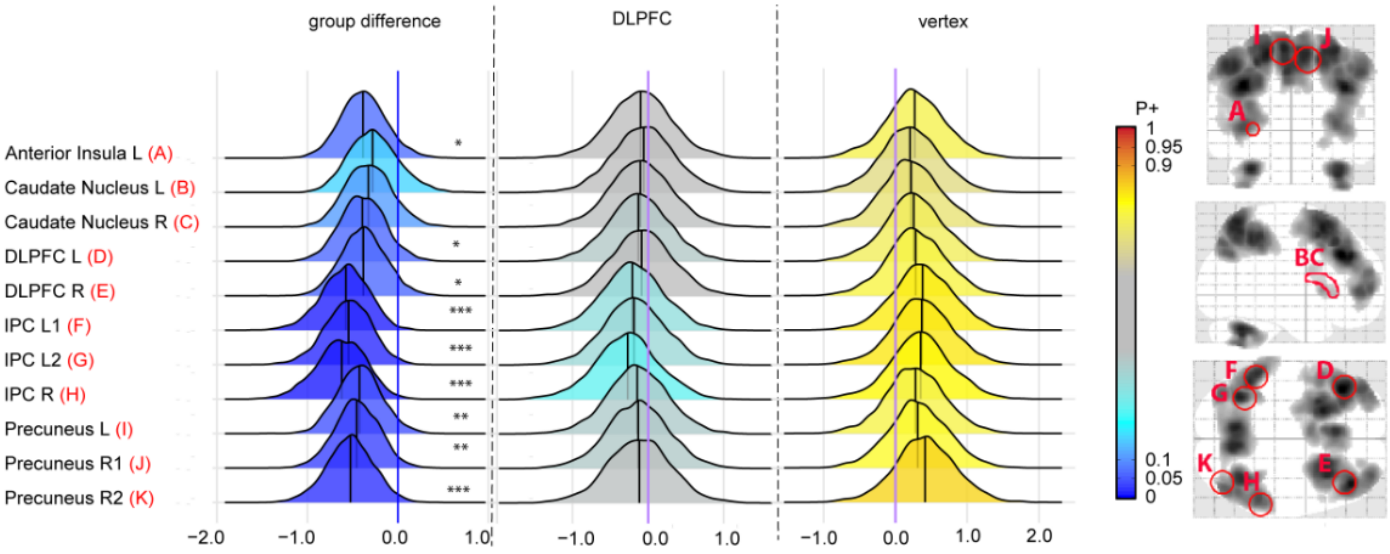

Figure S2: Whole-brain activation, TOL planning

Group differences (DLPFC vs vertex) in change in activation over time (T0 vs T2) at p<0.001, uncorrected. All clusters indicate a relative decrease in the DLPFC group in comparison to the vertex group. During the planning contrast, there was a decrease in activation following treatment in anterior and dorsolateral PFC, preSMA, posterior cingulate cortex, and cerebellum, as well as white matter in the corpus callosum , in comparison to the vertex group (Figure S5).

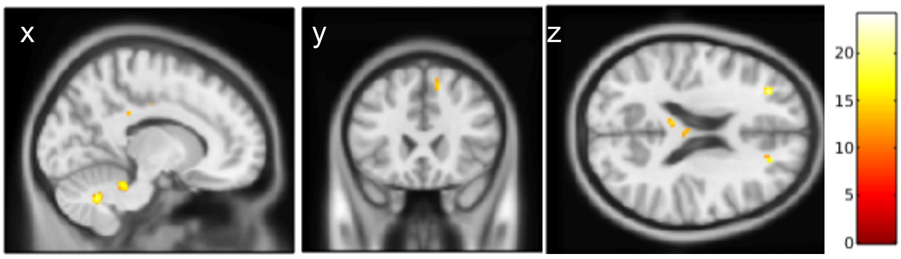


| **Brain region** | **Hemisphere** | **Cluster size (k)** | **MNI coordinates** | **Peak Z value** |
|---|---|---|---|---|
| Anterior prefrontal cortex | L | 23 | -32 / 40 / 20 | 4.34 |
| DLPFC (white matter) | R | 13 | 20 / 40 / 20 | 3.69 |
| Pre-supplementary motor area | R | 29 | 12 / 24 / 52 | 3.48 |
| Corpus callosum | L | 126 | -4 / -20 / 24 | 4.15 |
| Posterior cingulate cortex | L | 10 | -10 / -18 / 38 | 3.38 |
| Cerebellum | L | 14 | -26 / - 42 / -34 | 3.48 |
| Cerebellum | L | 68 | -10 / -60 / -36 | 3.99 |
| Cerebellum | L | 31 | -10 / -40 / -26 | 3.85 |
| Cerebellum | L | 11 | -20 / -50 / -40 | 3.33 |

Figure S3: Whole-brain activation, TOL taskload

Group differences (DLPFC vs vertex) in change in activation over time (T0 vs T2) at $p<0.001$, uncorrected. All clusters indicate a relative decrease in the DLPFC group in comparison to the vertex group. During the taskload contrast, the DLPFC group showed, in comparison to the vertex group, a decrease in activation in the inferior frontal gyrus, orbitofrontal white matter, frontal eye fields, insula, supplementary motor area, somatosensory cortex, ventral posterior cingulate, and supramarginal gyrus (Figure S6).

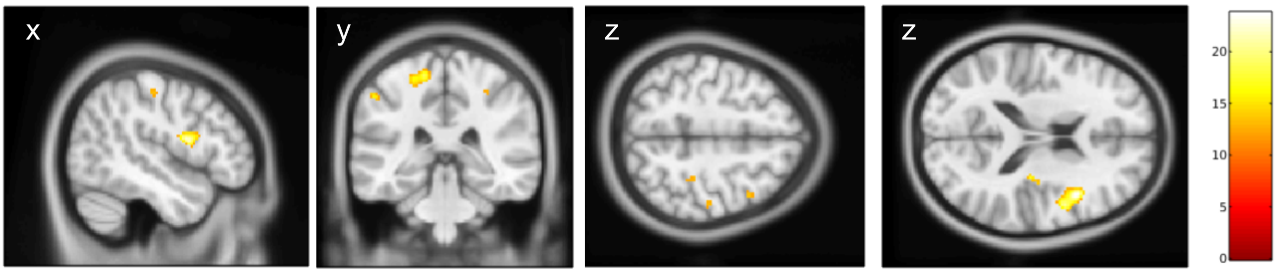


| **Brain region** | **Hemisphere** | **Cluster size (k)** | **MNI coordinates** | **Peak Z value** |
|---|---|---|---|---|
| Inferior frontal gyrus (opercular) | R | 195 | 42 / 12 / 18 | 4.31 |
| Orbitofrontal white matter | R | 30 | 24 / 34 / -4 | 4.13 |
| Frontal eye fields | R | 12 | 42 / 10 / 52 | 3.29 |
| Insula | R | 15 | 30 / -22 / 18 | 3.64 |
| Premotor/supple mentary motor | L | 17 | -12/ -4 / 60 | 3.72 |
| Somatosensory | L | 101 | -14 / -36 / 64 | 3.88 |
| Somatosensory | R | 15 | 48 / -20 / 50 | 3.25 |
| Somatosensory | R | 11 | 32 / - 32 / 50 | 3.25 |
| Ventral posterior cingulate | L | 39 | -10 / -16 / 36 | 3.72 |
| Supramarginal gyrus | L | 29 | -54 / -36 / 50 | 3.55 |
| Supramarginal gyrus | R | 11 | 56 / -28 / 48 | 3.27 |

Figure S4: SST response inhibition

Group differences (preSMA vs vertex) in change in activation over time T0 vs T2) at $p<0.001$, uncorrected. All clusters indicate a relative decrease in the preSMA group in comparison to the vertex group. During the response inhibition contrast, the preSMA-stimulated group showed, in comparison to the vertex group, a decrease in activation in the thalamus and frontal white matter following treatment (Figure S7).

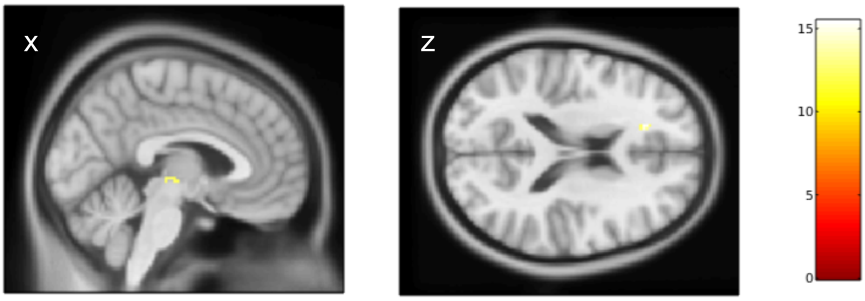


| **Brain region** | **Hemisphere** | **Cluster size (k)** | **MNI coordinates** | **Peak Z value** |
|---|---|---|---|---|
| Dorsal anterior cingulate cortex/ Frontal white matter | L | 10 | -18 / 32 / 18 | 3.54 |
| Thalamus | L | 12 | -6 / -22 / -2 | 3.25 |

Figure S5: SST error processing

Group differences (preSMA vs vertex) in change in activation over time (T0 vs T2) at p<0.001, uncorrected. All clusters indicate a relative decrease in the preSMA group in comparison to the vertex group. During the error processing contrast, the preSMA-stimulated group showed, relative to the vertex group, a decrease in SMA activation during error processing after treatment (Figure S8).

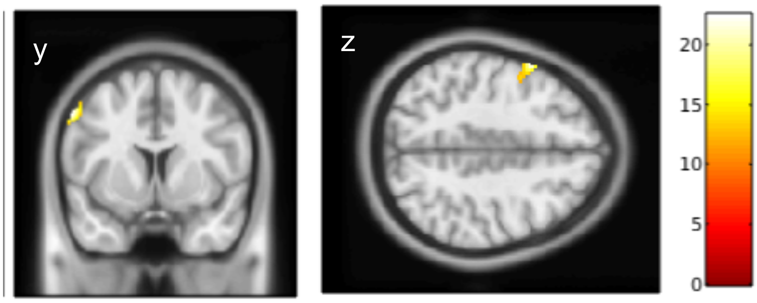


| **Brain region** | **Hemisphere** | **Cluster size (k)** | **MNI coordinates** | **Peak Z value** |
|---|---|---|---|---|
| SMA | L | 143 | -58 / 8 / 44 | 4.25 |

Fig. S6 : association between T0-T2 change in TOL planning activation and change in YBOCS for all participants: Posterior distributions show association between T0-T2 change in YBOCS and change in activation during planning for all participants combined (n=61). Evidence strength is visualized by the color bar, red indicating positive associations and blue negative. For interpretation we added ***= for very strong evidence (P+>0.975 or <0.025), **=strong evidence ((P+>0.95 or <0.05), or *=moderate evidence (P+> 0.90 or <0.10). Corresponding P+ values can be found in Table S11. Abbreviations: DLPFC, dorsolateral prefrontal cortex; IPC, inferior parietal lobe; L, left; R, Right YBOCS, Yale-Brown Obsessive-compulsive scale; ROI, region of interest

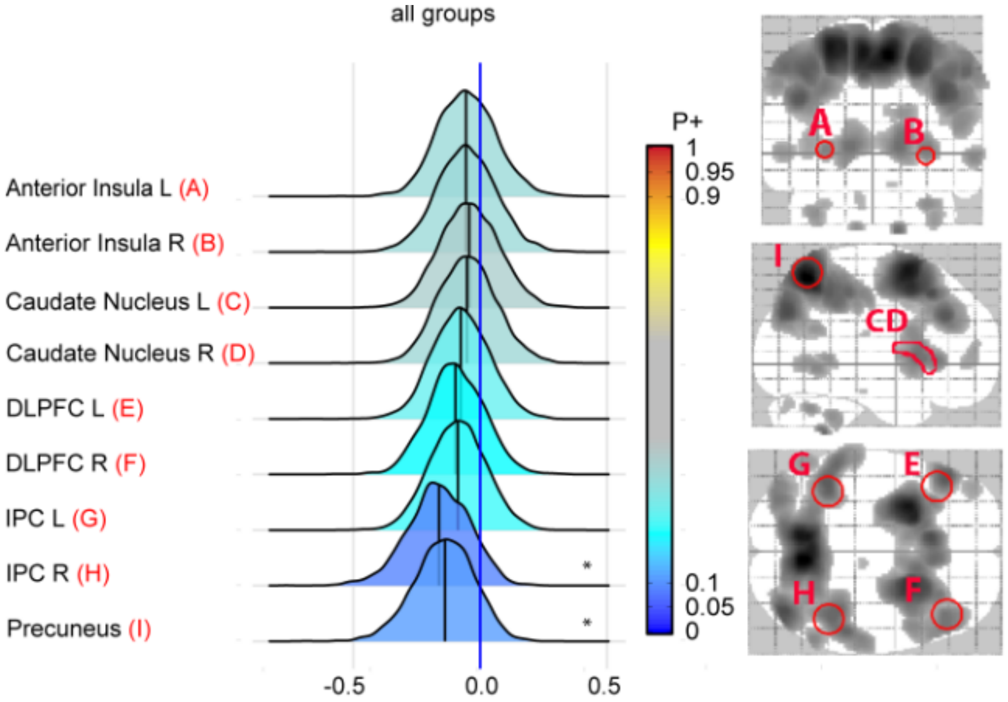

Figure S7: association between T0-T2 change in TOL taskload activation and change in YBOCS for all participants: Posterior distributions show association between T0-T2 change in YBOCS and change in activation during taskload for all participants combined (n=61). Evidence strength is visualized by the color bar, red indicating positive associations and blue negative. For interpretation we added ***= for very strong evidence (P+>0.975 or <0.025), **=strong evidence ((P+>0.95 or <0.05), or *=moderate evidence (P+> 0.90 or <0.10). Corresponding P+ values can be found in Table S11. Abbreviations: DLPFC, dorsolateral prefrontal cortex; IPC, inferior parietal lobe; L, left; R, Right; YBOCS, Yale-Brown Obsessive-compulsive scale; ROI, region of interest

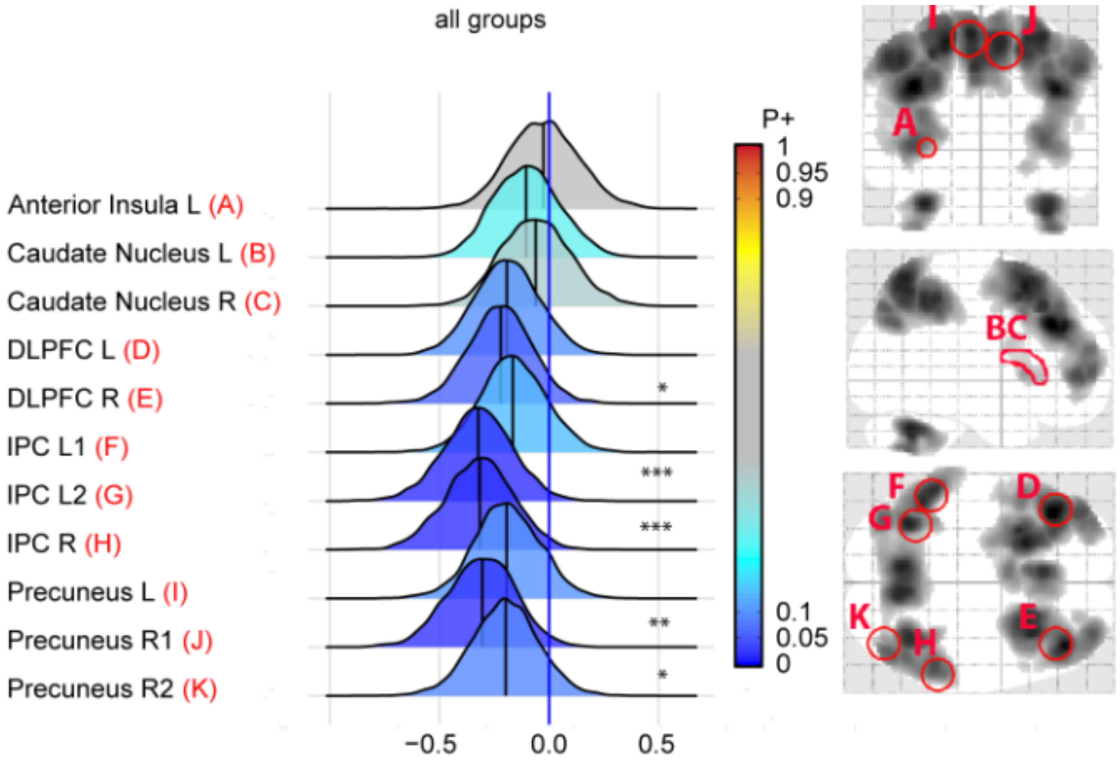
